# Within-host models unravelling the dynamics of dengue reinfections

**DOI:** 10.1101/2023.09.21.23295910

**Authors:** Vizda Anam, Bruno V. Guerrero, Akhil Kumar Srivastav, Nico Stollenwerk, Maíra Aguiar

## Abstract

Dengue fever is a major public health concern in tropical regions, caused by four distinct serotypes. Sequential infection with a different serotype increases the risks of severe disease through antibody-dependent enhancement (ADE). Huge modeling efforts have focused on primary and heterologous secondary infections, while the dynamics of homologous secondary infections were overlooked due to the assumption of lifelong immunity preventing reinfections by the same serotype.

Recent findings challenge the current understanding of dengue immunity. To explore immunological responses in various dengue infection scenarios, we use a within-host modeling framework that considers individual immunological variations. These models are validated using empirical data. In addition to successfully capturing primary and heterologous secondary infection dynamics facilitated by ADE, this framework provides, for the first time, insights into homotypic reinfection dynamics and discusses its relevance in dengue transmission at the population level, with potential implications for disease prevention and control strategies.

## 1 Introduction

With more than one-third of the world’s population at risk of infection, dengue fever is a major public health problem in the tropics and subtropics [1]. Caused by four antigenically related but distinct serotypes, DENV1, DENV2, DENV3 and DENV4, it was believed that an infection with one serotype would grant lifelong immunity to that specific serotype, effectively preventing any subsequent infections by the same serotype (homologous infection). However, recent large cohort studies have shown that homologous secondary infections may not always result in complete protection against re-infection [2]. In the study by four patients experiencing homotypic dengue reinfections being identified, challeging the current understanding of dengue immunity.

Nonetheless, temporary cross-immunity (TCI) to other serotypes exist. In combination with the seasonal patterns common to vector-borne diseases, TCI leads to a temporal gap between primary and secondary infections, shaping dengue transmission dynamics at population level.

Most of the primary infection cases recover with non-severe clinical symptoms, and treatment for uncomplicated cases is primarily supportive. There is, however, good evidence that sequential infection with a different serotype (heterologous infection) increases the risk of developing severe disease due to a process described as antibody-dependent enhancement (ADE) [3, 4, 5, 6, 7, 8]. ADE occurs when pre-existing antibodies from a previous dengue infection do not neutralize but rather enhance the new infection.

A safe, effective and affordable dengue vaccine against the four serotypes would represent a significant advance for reducing disease transmission. As of now, two tetravalent dengue vaccines have completed the phase 3 clinical trial. Dengvaxia, developed by Sanofi Pasteur, now licensed in several countries [9], and the Qdenga (TAK-003) vaccine, developed by Takeda Pharmaceutical Company [10, 11], recently received approval in Indonesia, Brazil, and the European Union.

It is noteworthy to mention that Dengvaxia has resulted in serious adverse events in vaccinated individuals without previous exposure to any of the dengue serotypes (seronegative) compared with age-matched seronegative controls [12, 13, 14, 15, 16, 17]. Because of that, its use is now restricted to individuals with confirmed previous dengue infection (seropositive). On the other hand, Qdenga vaccine has shown higher and more balanced vaccine efficacy against virologically confirmed dengue disease and hospitalization than efficacies reported for Dengvaxia [18]. Nevertheless, long-term surveillance consisting of cautious observation of Qdenga vaccine recipients is required [19, 18, 20], since serotype-specific negative vaccine efficacy was also observed for vaccinated seronegative individuals [21].

Mathematical modeling offers a powerful tool for studying disease dynamics at different scales. At the individual level (within-host), models can help understanding the immune response dynamics, the impact of previous infections on disease severity, and the effectiveness of interventions such as vaccines or antiviral treatments. At the community level, mathematical models can predict disease transmission based on the serological population profile considering biological parameter values estimated from incidence data. A valid framework will be used to evaluate the effectiveness of vector control strategies, such as insecticide spraying or eliminating mosquito breeding sites, and vaccination in reducing disease transmission, for example.

Huge modeling efforts focusing on various aspects of dengue dynamics, including immunological and epidemiological factors, been performed in the past 10 years [22, 23].

Within-host modeling approach is built to describe viral replication and immunological responses during dengue infection processes Frameworks such as those discussed in [24, 25, 26, 27, 28, 29, 30, 31, 32, 31, 33, 34], have considered the dynamic interaction between free virus and susceptible target cells, differing in the functional form to model viral infectivity, immune response mediated by antibodies, and/or viral clearance dynamics.

In most cases, model parameters are estimated from statistical data analysis derived from cohorts evaluating data on human viremia and infected mosquitoes, providing insights on transmission parameters such as the rate of transmission from infected humans to mosquitoes [35]. Up to date, only few modeling studies have used empirical immunological data for direct model parametrization and validation. To cite some of them, the models proposed by Clapham et al. in [36, 27] and by Ben-Shachar et al. in [25] which were parameterized by fitting patient data on antibody titers and/or viral load measurements recorded sequentially during infection. These data were obtained from a clinical trial conducted by Nguyen et al. [37].

Based on immunological indicators such as the magnitude and time of peak viraemia, these models have focused on understanding the antibody-specific dynamics during the infection clearance process of primary infection and/or the increased risk of severe disease associated with secondary heterologous infections. However, the immunological dynamics of homologous secondary infections have been consistently overlooked. This neglect was based on the belief that an initial infection with one serotype would confer lifelong immunity to that serotype, effectively preventing any subsequent infections by the same serotype. As a result, dengue transmission models have never taken into consideration the potential impact of homologous infections in a population.

With advances on laboratory tests, a recent large cohort study conducted by Waggoner et al. [2] has found that immunity to homologous serotypes may not always be fully sterilizing, with patients experiencing homologous reinfections being identified. The extent of dengue transmission caused by homologous infection remains largely unexplored, highlighting the need for further research to understand the impact and implications of these reinfections on the overall dengue dynamics.

In this paper, we introduce a novel modeling framework that considers individual immunological variations, able to describe well the dynamics of primary and secondary dengue infections. The models are parametrized using data on viral load and antibody concentrations on patients experiencing dengue infections with all four serotypes, as reported from the clinical trial of the drug balapiravir to treat dengue infection [37]. Building upon previous studies [27, 25], which use immunological data on patients experiencing dengue infections with serotype 1 (DENV1) and serotype 2 (DENV2), our analysis extends to include, for the first time, data on DENV3 and DENV4 infections.

Using a within-host modeling framework accounting for the inherent individual immunological variances during the infection process, we aim at understanding the characteristics of immunological responses during primary and secondary infections. Special attention is given to the dynamics of homotypic secondary infection, an aspect that has not been unexplored in research modeling.

## Results

### The available immunological data

For the modeling framework parametrization we use the immunological data consisting of viral load measurements as well as IgM and IgG antibody titters. The data were obtained from a clinical trial evaluating the efficacy of the drug balapiravir in treating dengue infection, as published in Nguyen et al. [37], publicly available from the study by Clapham et al. [27].

The trial included a cohort of sixty four (64) patients infected with dengue, with thirty two (32) individuals identified with DENV1 infection, twenty one (21) with DENV2, five (5) with DENV3, and six(6) with DENV4. While data on DENV1 and DENV2 infections were used in previous studies [27, 25], here we present the first analysis of the data on DENV3 and DENV4 infections.

According to the study by Nguyen et al. [37], viral load measurements were taken twice a day for all patients. IgM and IgG antibody titers were measured throughout the infection period using an ELISA assay, with quantification based on optical density. Among the sixty four (64) patients with confirmed dengue infection, being five (5) cases reported as primary infections, while the rest of the cases were classified as secondary infections. However, this study did not provide any additional information regarding the serotype responsible for these primary and secondary infections.

It is worth mentioning that the administration of balapiravir treatment did not result in measurable changes in various virological, clinical, or immunological endpoints of the trial study. Therefore, in this study, all the available data, including samples from placebo treatment and no treatment groups, were used for parameterizing the models.

### Data preparation process for model calibration

As the modeling framework does not incorporate serotype structure, the serotype-specific data is treated as individual data sets. In other words, we consider four separate data sets, each corresponding to the infections caused by one of the four dengue serotypes.

The data preparation process for modelling parametrization consisted of two steps:

1. We have performed a thorough data inspection, plotting the serotype dengue specific viral (measured RNA copies/ml) and antibody (quantified via measurement of optical density) titters over time for each patient. The limit of virus detection was determined based on the findings reported by Nguyen et al. [37], with 357 copies/mL for DENV1 and DENV3, and 72 copies/mL for DENV2 and DENV4, while the limit of antibodies detection was established according to Clapham et al. [27], with 25 optical density units as maximum level for both anybody types.
2. For each serotype-specific data set, individuals exhibiting similar immunological features in terms of viral and antibody concentration levels were categorized according to their type of infection: primary infection, homologous secondary infection, and heterologous secondary infection.

The classification of subjects was performed based on the immunological features described in the literature, as follows:

A. **Primary infection (Group 1):** subjects with viral titers and IgM antibody concentration levels above the detection threshold, and with IgG concentration below the detection threshold.
B. **Secondary homologous infection (Group 2):** subjects with viral titers below the detection threshold, and both IgG and IgM antibody levels above the detection threshold.
C. **Secondary heterologous infection (Group 3):** subjects with viral titers and both IgG and IgM antibody levels above the detection threshold.

This classification is shown in Fig. 1 for DENV1 subjects and in Fig. 2 for DENV2, DENV3, and DENV4 subjects. It is worth noting that the classification of secondary homologous infection based on immunological features (highlighted in dark blue) is presented here for the first time.

**Figure 1:**
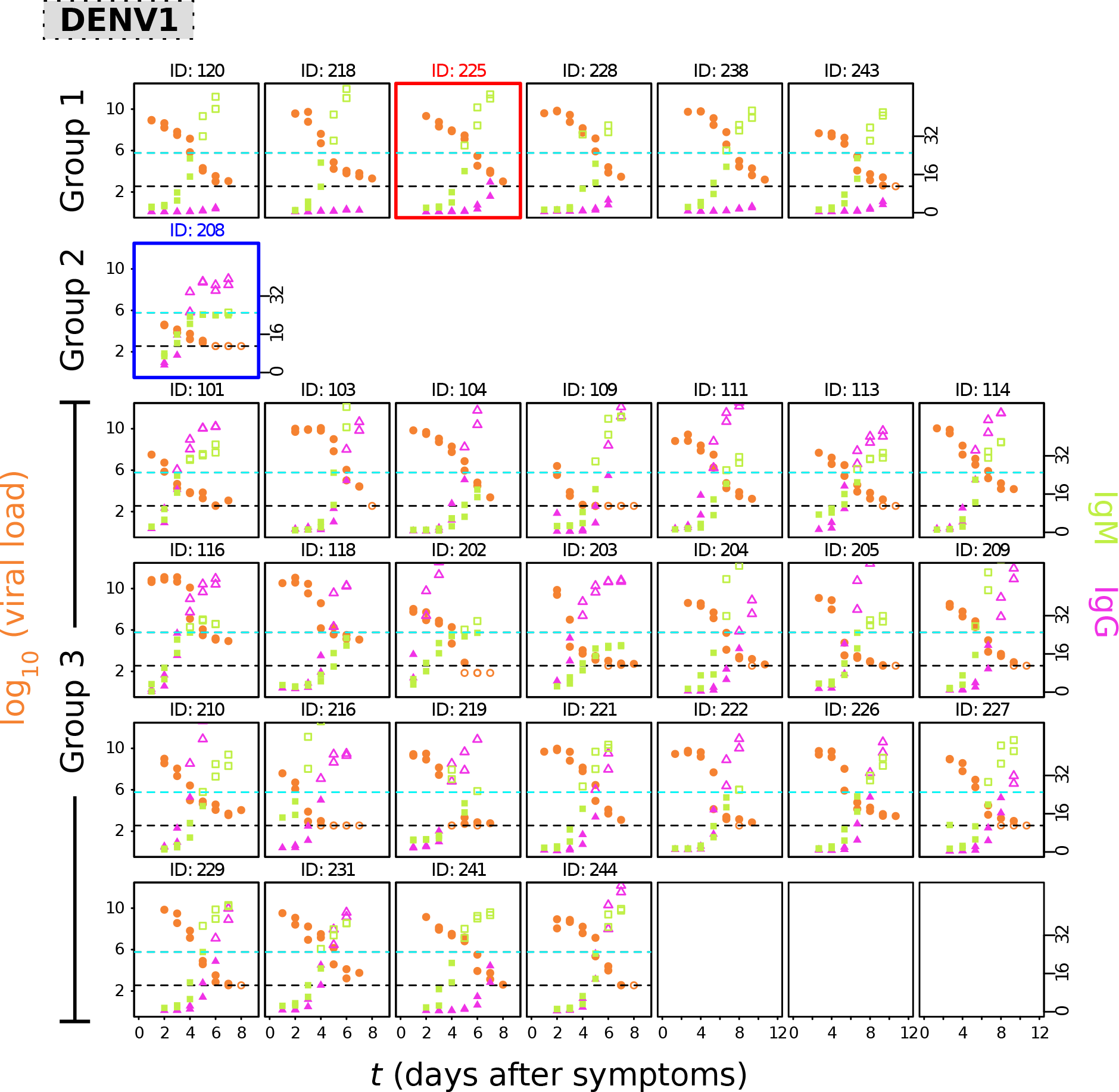
The DENV1 data set. For subjects infected with DENV-1, each box in the figures represents an individual sample. The viremia levels, measured in log10 RNA copies/mL of plasma, are represented by orange dots. The IgM and IgG antibody titters, measured using optical density and plotted on a linear scale, are depicted by green squares and magenta triangles, respectively. For accurate detection of antibodies, a maximum level of 25 optical density units was set, as shown by the dashed cyan lines. The limit of virus detection falls between 1500 copies/mL and 15000 copies/mL, as indicated by the dashed black lines. Unfilled marker symbols show measurements below the assay limit of detection for virus and above the upper limit of reliable (linear) quantification for IgG and IgM. Highlighted samples represent cases where the classification diverges from previous studies and will be subjected to separate analysis using our modeling framework.

**Figure 2:**
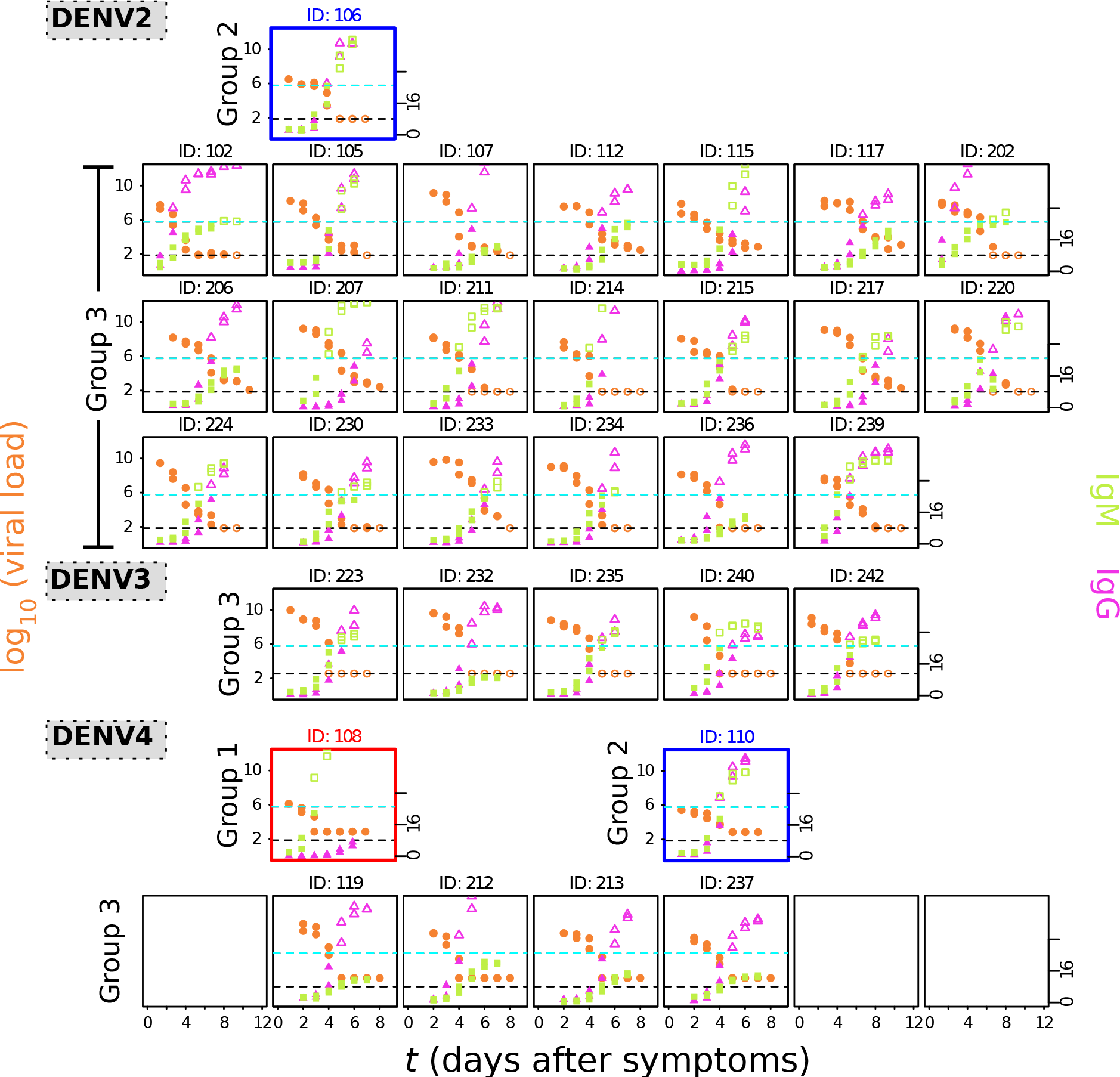
The DENV2, DENV3 and DENV4 data sets. For subjects infected with DENV2, DENV3, and DENV4, each box in the figure represents one infected individual with a specific serotype. The viremia levels, measured in log10 RNA copies/mL of plasma, are represented by orange dots. The IgM and IgG antibody titters, measured using optical density and plotted on a linear scale, are depicted by green squares and magenta triangles, respectively. For accurate detection of antibodies, a maximum level of 25 optical density units was set, as shown by the dashed cyan lines. The limit of virus detection falls between 1500 copies/mL and 15000 copies/mL, as indicated by the dashed black lines. Unfilled marker symbols show measurements below the assay limit of detection for virus and above the upper limit of reliable (linear) quantification for IgG and IgM. Highlighted samples represent cases where the classification diverges from previous studies and will be subjected to separate analysis using our modeling framework.

For the DENV1 data set, Fig. 1, we have identified six patients experiencing a primary dengue infection, and twenty-five patients experiencing secondary dengue infection. In contrast to the classification given in [27], three patients have been reclassified upon a careful data inspection.

Patient ID: 225 (highlighted in red), previously treated as secondary heterologous infection, is now reclassified as a primary infection, as the sample presents high viral titters, high IgM antibody concentrations, and low IgG antibody concentrations. We note, however, that in this case, the IgG concentration is just at the threshold of detection. Given that the trial was conducted back in 2013, this IgG level could have made accurate classification challenging and possibly resulted in unspecific serological outcome.

Patient ID: 208 (highlighted in blue), previously excluded from the analysis conducted in [27], has been classified here as a secondary homologous infection, as this sample presents relatively low viral titter (as compared to the other samples in the same data set), along with high IgG antibody concentrations, and lower IgM antibody concentration levels.

For the DENV2 data set, Fig. 2, only secondary infections were identified. However, Patient ID: 106 (highlighted in blue), initially treated as secondary heterologous infection in [27], has been reclassified as secondary homologous infection, supported by the patient’s low viremia levels and high levels of IgG and IgM antibodies.

Furthermore, in this study we show the first-time analysis of data for patients with DENV3 and DENV4 infections. In the DENV3 data set, only secondary heterologous infections were identified. As for the DENV4 data set, subjects were classified as follows:

Patient ID: 108 (highlighted in red) was classified as primary infection. This sample presents moderate viral load and high IgM levels, while the concentration of IgG antibodies is observed to be near the threshold of detection, with possible unspecific serological result.

Patient ID: 110 (highlighted in blue) was classified as secondary homologous infection. The basis for this classification is the observation of low viremia and high IgG and IgM concentration levels in the patient’s sample.

The remaining four subjects in this data set were classified as secondary heterologous infections. These patients exhibit high viremia and high IgG concentration levels, along with lower IgM concentration levels.

### Data classification deviating from prior studies

Our analysis of the data from the study conducted by Nguyen et al. [37] has shown discrepancies in the classifications used in previous modeling studies. These inconsistencies introduce uncertainties and emphasize the need for special attention during the data interpretation process.

In contrast to the data categorization used in the modeling studies by Clapham et al [27] and by Ben-Shachar et al. [25], which agrees with the reported number primary and secondary infections in Nguyen et al. [37], our analysis has identifies a total of 7 (instead of 5) subjects experiencing a primary dengue infection. These additional two primary infections, namely patient ID DENV1-225 and patient ID DENV4-108, have showed IgG concentration levels near the threshold point, which may have resulted in unspecific serological results. These samples exhibited significantly higher concentrations of IgM, which align closely with patterns typically observed in primary infection cases.

Additionally, from our analysis of the secondary infection cases, patients ID DENV1-208, ID DENV2-106, and ID DENV4-110 were classified as homologous secondary infection. This novel classification, discussed here for the first time, has not been previously considered in modeling studies.

Considering the uncertainties in the data classification process, we further investigated these samples using our models. The results of this analysis are presented in the Supplementary Material, shown in Figures S1-S5, supporting the current classification of these cases.

### Modeling dengue infection process: simplified description of complicated dynamics

During a dengue infection, the presence of RNA copies of the virus in a blood sample allow to identify the specific serotype causing the infection. However, to distinguish between primary and secondary infections, it is essential to consider both viremia levels and antibody dynamics together.

The left-hand side of Figure 3, a comprehensive representation of the development of a primary dengue infection is illustrated and matched with the model simulation. Vertical solid lines are used to align these phases with the Model A simulation (described in detail in the nest subsection), which is shown in the lower left side of the figure. Orange line refer to the viremia dynamics, whereas green and magenta lines represent the antibodies in response to the infection. Free antibodies and virus-antibody complexes are plotted with solid and dashed lines respectively.

**Figure 3:**
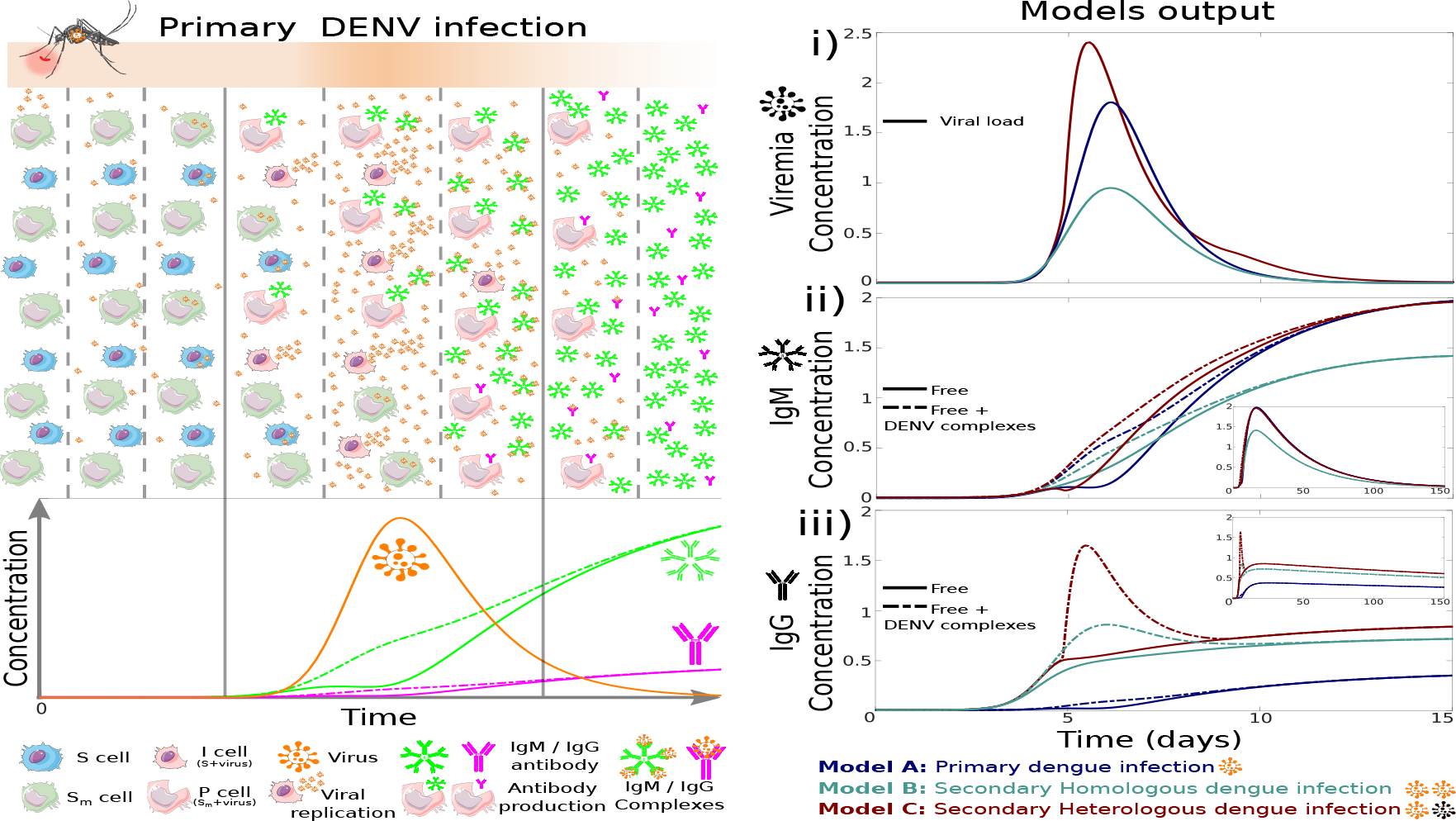
Left-hand side: the schematic representation of a primary dengue infection process is illustrated in the upper part, matching the immunological dynamics obtained through mathematical modeling (Model A) in the lower part. Right-hand side: modeling outputs for the dynamics of viral load (i) and IgM (ii) and IgG (iii) antibodies are shown for primary and secondary, homologous and heterologous, infections.

The right-hand side of Fig. 3 presents a comparative modeling outputs for primary and secondary dengue infection dynamics for the viremia and antibody levels (IgM and IgG) over time. Subfigure (i) shows the evolution of viremia throughout the course of each infection type. In a similar way, the dynamics of IgM and IgG antibodies are shown in subfigures (ii) and (iii), respectively.

In the Methods section, we introduce the baseline modeling framework, Model A, which describes the dynamics of a primary dengue infection. Model A is extended to describe the dynamics of secondary dengue infections, considering the pre-existing serotype-specific antibodies generated during a primary infection. Homologous reinfection is modeled with Model B while heretologous reinfection is modeled Model C respectively.

By examining these dynamics separately, we gain a better understanding of the patterns of viral replication within the host during the different stages of the disease progression and how the immune system responds to the presence of the virus through the production of antibodies. The latter is essential for comprehending the development of immune memory providing protection against subsequent infections.

### Modeling primary dengue infections

Primary dengue infection occurs when an individual is exposed to one of the four dengue virus for the first time. The viral replication process leads to an increase in viremia level, and as the infection progresses, the immune system begins producing antibodies. These antibody levels gradually rise and play a crucial role in clearing the infection, conferring a certain degree of immunity to subsequent infections.

This process can be explained in detail with the help of the upper left side of Figure 3, showing the virus transmission to a human host by an infected female Aedes mosquito. The color intensity of the orange band reflects in a phenomenological way the viremia in the human host over time. The schematic representation of the primary dengue infection process is divided into three blocks, with each block representing a specific phase of the infection process. These blocks are further subdivided into eight columns, providing a detailed breakdown of the different stages within each phase. In detail, free viral particles infecting target cells leads to viral replication (block 1, columns 1-3). As the infection progresses, the immune system begins producing antibodies, first the IgM (then the IgG in a much lower scale), as a response to the virus replication (block 2, columns 4-6). The antibodies bind to the free virus, creating virus-antibody complexes, clearing the infection (block 3, columns 7-8).

The primary infection immunological process is accurately described by the deterministic model, as illustrated in the lower left side of Figure 3. For a comprehensive overview of the deterministic modeling outputs, please refer to the works of Sebayang et al. [33] and Anam et al. [34].

In this study, we use both the deterministic and the stochastic approaches to describe the dynamics of the infection process. As shown in Figure 4, numerical simulations of Model A are compared to the immunological data obtained from individuals who have experienced primary infection with DENV1 and DENV4. The deterministic solutions (navy lines) describes the patterns observed in the immunological data, whereas the 500 stochastic realizations (grey lines) show the extent to which the outcomes of random individual realizations are anticipated to deviate from the expected average solution (e.g. mean field solution). For both date sets, the confidence band given by the stochastic simulations contain all the real patients’ immunological data, describing well the immunological dynamics of primary infections.

**Figure 4:**
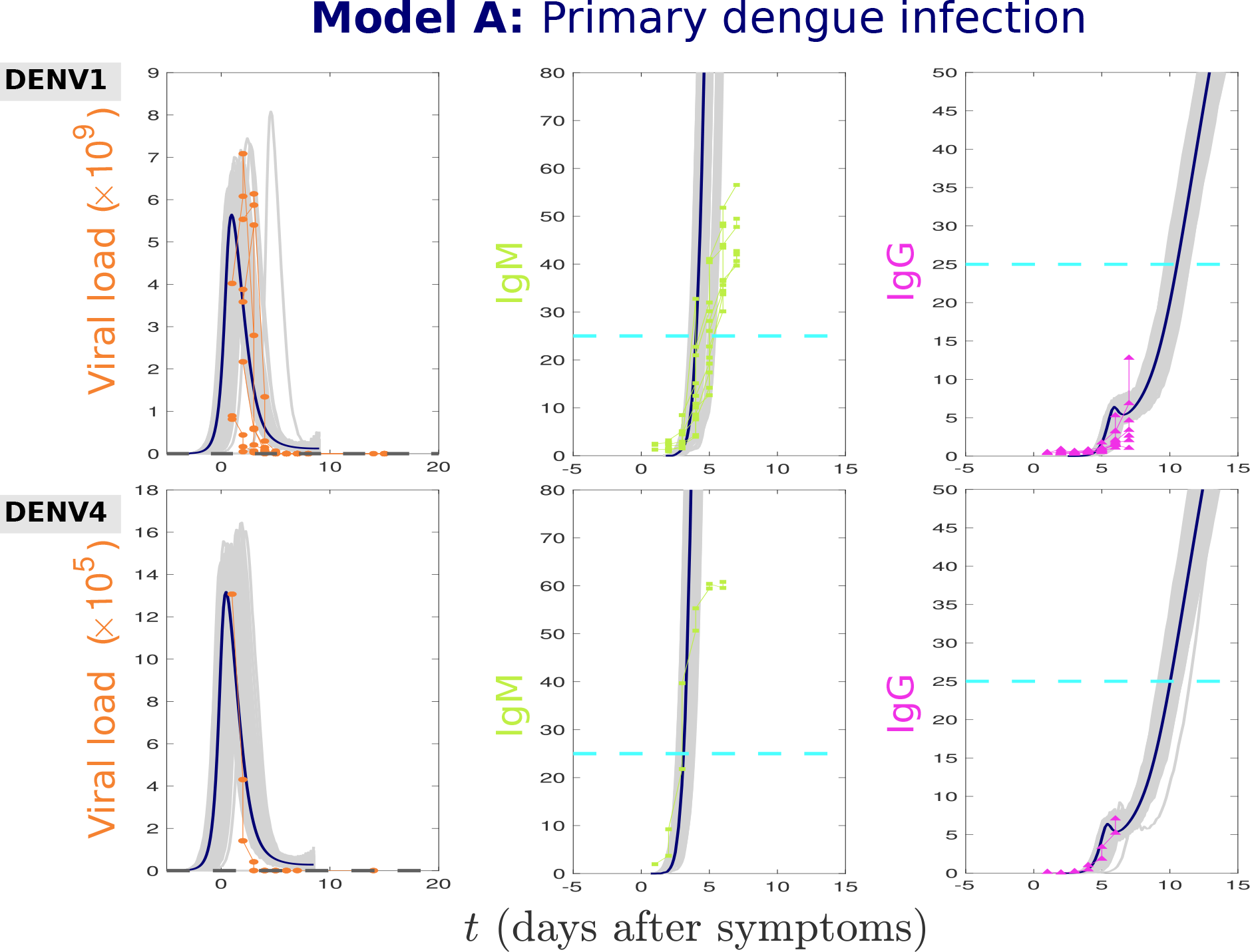
Stochastic realizations (grey line) and the deterministic (navy solid line) solution of the Model A are compared to the immunological data obtained from individuals experiencing primary infection with DENV1 and DENV4 serotypes. Viral load is shown in orange, whereas antibody concentrations are shown in green (IgM) and in magenta (IgG). The parameter values used for modeling simulations are shown in Table S1 available in the Supplementary Material.

Note that the data follows the same color convention used in the model simulation shown in the lower left-hand side of figure 3: orange for viral load, and green and magenta for IgM and IgG antibodies concentrations, respectively.

### Modeling secondary dengue infections

After a primary dengue infection, the initial high levels of cross-reactive antibodies offer some protection against reinfection by any dengue serotypes. Over time, these antibody levels gradually decline. The IgM antibody level can remain detectable in the organism for a short period (around 12 weeks post symptom onset), while the serotype-specific IgG antibody persists for a longer duration (many months and even years after a dengue infection). As immunity wanes, individuals become susceptible to acquiring a secondary infection.

Initial studies have suggested that a dengue infection could provide lifelong sterilizing immunity against a specific serotype, effectively protecting individuals from future reinfections with that same serotype. As a result, mathematical dengue models have predominantly centered on understanding and describing the dynamics of primary and secondary heterologous infections, often overlooking the dynamics of homologous secondary infections. Nonetheless, the occurrence of homologous secondary infections reported in a recent trial by Waggoner et al. [2], has challenged the current understanding of dengue immunity. These findings have important implications for modeling dengue transmission.

In this study, we introduce a novel model that, for the first time, presents the dynamics of homotypic reinfection. Accounting for an extra term to represent the pre-existing serotype-specific antibodies, the dynamics of secondary homologous infections are modeled using Model B. The heterologous infection dynamics account for extra terms to describe the antibody-dependent enhancement process and is modeled with Model C.

As with Model A, we use both deterministic and stochastic approaches, with the deterministic solutions represented by navy lines, and 500 stochastic realizations displayed as grey lines. The model simulations are compared to the immunological data obtained from individuals who have experienced secondary infections with all dengue serotypes, as shown in Figs. 5, for those in Group 2, and in Fig. 6, for those in Group 3.

**Figure 5:**
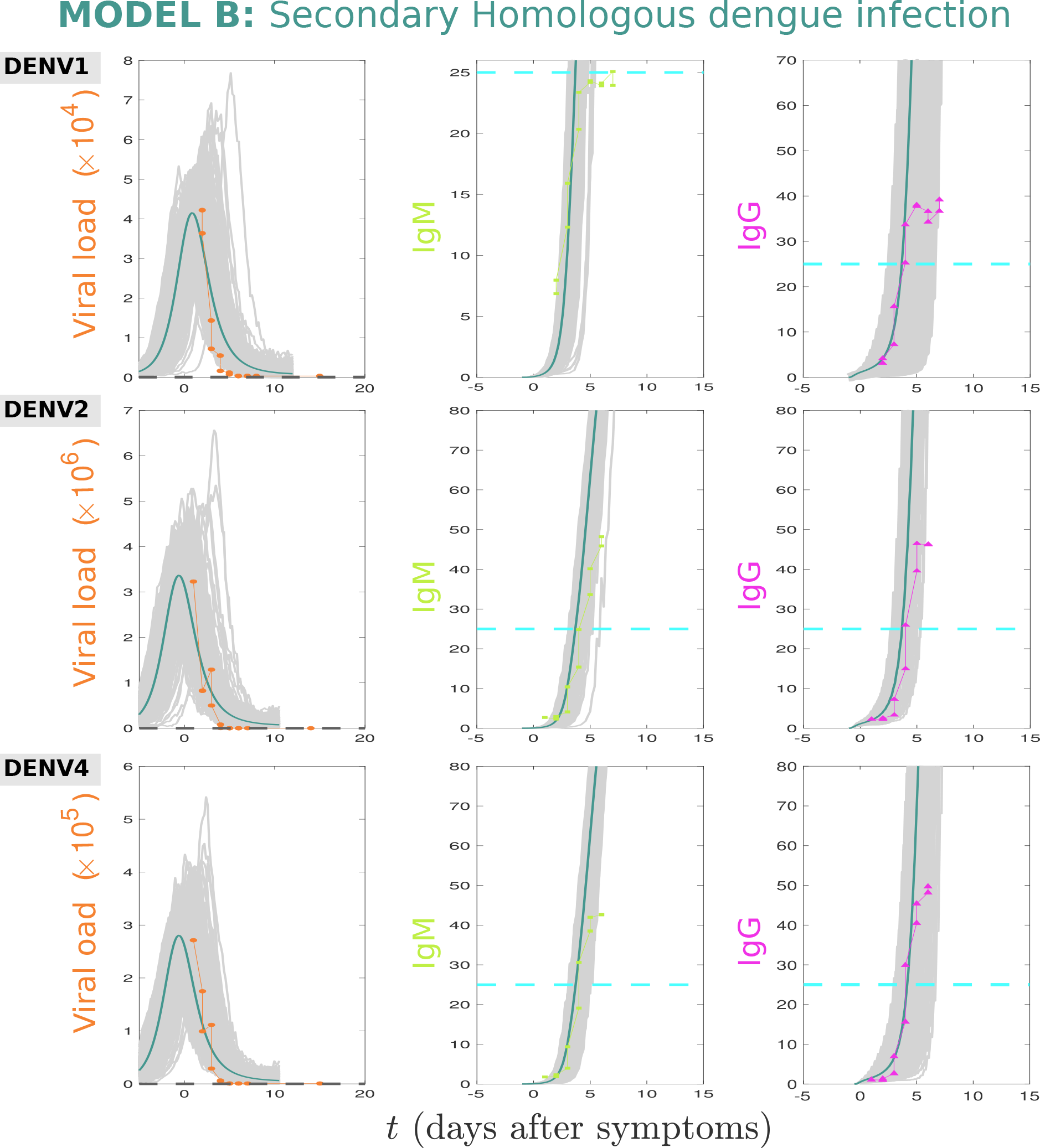
Stochastic realizations (grey line) and the deterministic (navy solid line) solution of the system are compared to the immunological data obtained from individuals who have experienced primary infection with DENV1, DENV2 and DENV4 serotypes. Viral load is shown in orange, whereas antibody concentrations are shown in green (IgM) and in magenta (IgG). The parameter values used for modeling simulations are shown in Table S1 available in the Supplementary Material.

**Figure 6:**
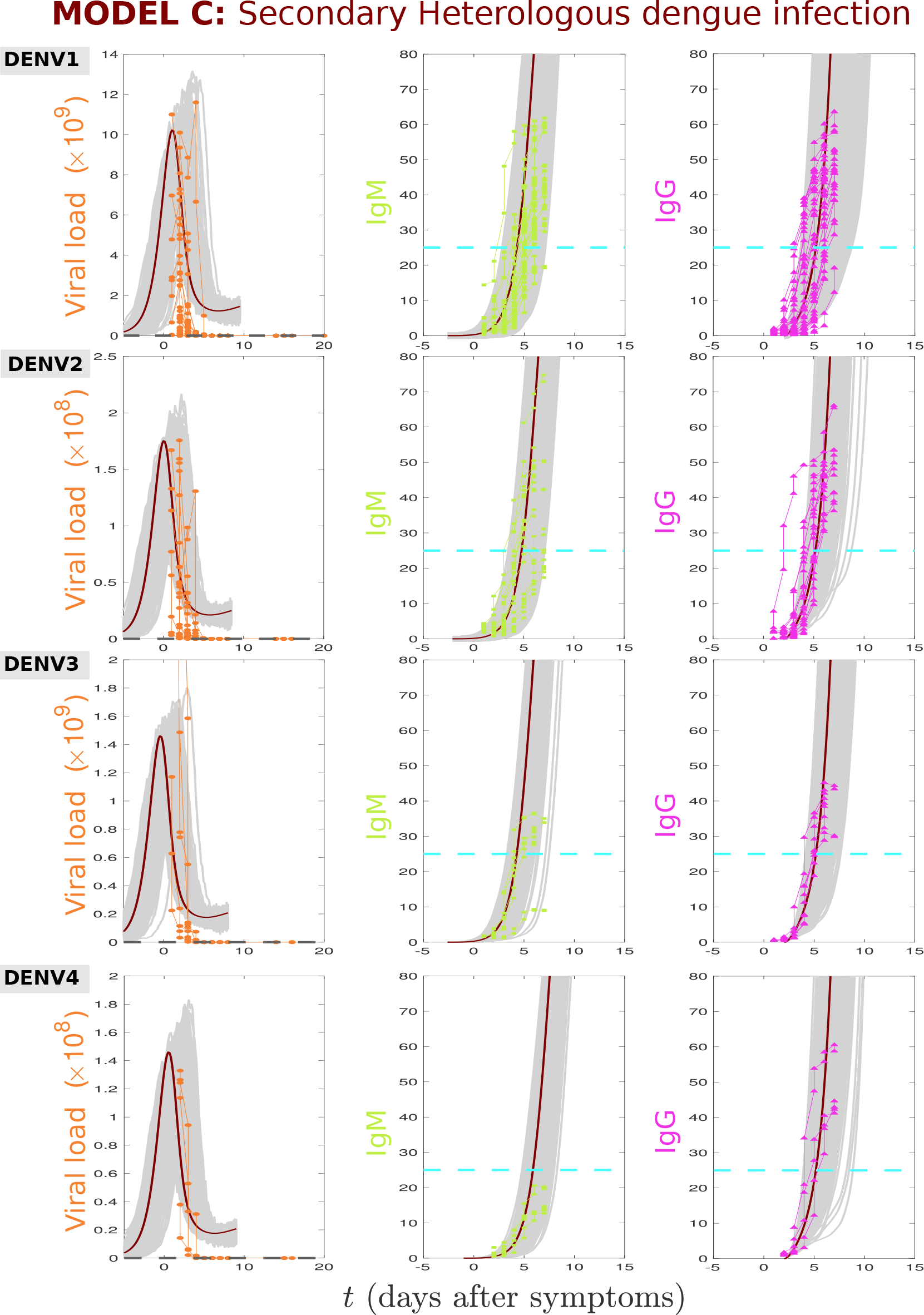
Stochastic realizations (grey line) and the deterministic (navy solid line) solution of the system are compared to the immunological data obtained from individuals who have experienced primary infection with DENV1, DENV2, DENV3 and DENV4 serotypes. Viral load is shown in orange, whereas antibody concentrations are shown in green (IgM) and in magenta (IgG). The parameter values used for modeling simulations are shown in Table S1 available in the Supplementary Material.

### Unravelling the immunological dynamics of homologous secondary dengue infections

The recent large cohort study carried out by Waggoner et. al. [2] has provided valuable insights into the duration and efficacy of immunity against homologous dengue reinfections. The authors have reported four patients with homotypic DENV reinfections. These patients developed viremia, with or without clinical symptoms, 1-2 years after a previous dengue infection caused by the same serotype. Their findings indicate that immunity to homologous serotypes may not always be fully sterilizing, suggesting that serotype-specific immunity to dengue may be short-lived. Considering this, it is crucial to carefully investigate the implications of such short-term immunity on disease transmission dynamics.

In its simplicity, the immunological dynamics of a secondary homologous infection can be described as follows.

Once the infection process is established, with free virus infecting target cells, presentation of viral antigens triggers the production of pre-existing serotype-specific IgG antibodies. These antibodies rapidly increase in concentration within a short period. These serotype-specific antibodies bind and effectively neutralize the homotypic virus, clearing the infection in a much faster scale than in a primary infection.

This rapid and effective response leads to a low viremia, with individuals mostly developing asymptomatic/mild clinical manifestation of the disease. The high levels of IgG are observed to play a major role during the clearance of the homologous secondary infection, as opposed to the process observed for a primary infection, in which the IgM antibody type is crucial for the infection clearance.

In Figure 5, we present a comparison between the numerical simulations of Model B and the immunological data from Group 2, referring to individuals experiencing secondary homologous infection (classified based on the patients’ low viremia levels and high levels of IgG and IgM antibodies).

The numerical simulations of Model B describe well the immunological data for DENV1, DENV2 and DENV4, reinforcing the validity of the model’s representation of secondary homologous infections.

### The dynamics of heterologous secondary dengue infections via the ADE process

In simple terms, the immunological dynamics of a secondary heterologous infection can be described as follows. Free virus particles invade target cells, initiating the infection process. During this stage, the presentation of viral antigens triggers the production of pre-existing serotype-specific IgG antibodies. These antibodies rapidly increase in concentration, however, in the case of a heterotypic virus (a different serotype), these pre-existing antibodies do not neutralize but rather enhance the new infection. As the viral replication continues, viremia levels can reach higher peaks, which has been shown to be correlated with disease severity [38, 39, 40, 41].

This phenomenon is known as antibody-dependent enhancement (ADE) [4, 5, 3, 6, 7]. There is good evidence that sequential infection with a heterologous serotype increases the risk of developing severe disease due to ADE process. In this context, high viremia leads to an intensified immunological response mediated by IgM antibodies, contributing to the development of hemorrhagic manifestations that can progress to shock and ultimately, death [5, 42, 43, 44, 45].

Similarly to the process observed for a primary infection, the high levels of IgM are observed to play a major role during the clearance of the heterologous secondary infection, however, in this case, both antibodies reach much higher levels compared to primary and secondary homologous infections.

In Figure 6, we present the comparison between the model simulations and the immunological data for all serotypes. Model C can describe well the data from Group 3, referring to individuals experiencing secondary heterologous infection (classified based on the patients’ high viremia levels and high levels of IgG and IgM antibodies).

## Discussion

Aiming at understanding the immunological responses during dengue infections, in this work we analyze and expand a within-host modeling framework. Deterministic models that describe the immunological dynamics of primary and secondary infections are extended to their stochastic counterparts, accounting for the inherent individual immunological variances during the infection process.

The models were validated with empirical immunological data on viral load and IgM and IgG antibody titers. Besides using data on DENV1 and DENV2, which were previously used to validate simple models, this study also includes, for the first time, data on DENV3 and DENV4 infections.

Our modeling framework has shown to be effective in capturing and describing the immunological data for each dengue infection scenario, with distinct patterns of immune response clearly observed. In the case of primary infection (Model A), we have demonstrated that the high levels of the IgM antibodies play a crucial role in clearing the infection. On the other hand, for the dynamics of homologous reinfection (Model B) we have demonstrated that the high levels of the pre-existing IgG antibodies are responsible for clearing the infection, which occurs in a much faster scale than in a primary infection. However, it is important to note that despite faster clearance, viremia is observed, indicating the potential for disease transmission at the population level. This scenario has not been previously explored through mathematical modeling, and the results fill a knowledge gap in the understanding of the complex immune dynamics in different dengue infection scenarios.

In contrast, in the case of secondary heterologous infection (Model C), we have shown that the pre-existing (serotype specific) IgG antibodies contribute to viremia augmentation, analogous to the ADE process. Moreover, the excessive levels of IgM antibodies produced in a later stage of this infection process may contribute to the development of more severe symptoms and complications during the course of the infection, as reported in previous studies.

The findings from this study significantly contribute to our understanding of the immune responses occurring during both primary and secondary infections. Adding valuable insights to the existing mathematical modeling research on dengue transmission, our results support the occurrence of reinfection by the same serotype and its role on dengue transmission at the population level must be further investigated.

## Methods

In this study, two distinct mathematical modeling approaches are used to investigate the immunological dynamics of dengue infection process. The first approach involved solving deterministic models, which provide the qualitative behavior of viral load and antibody dynamics during primary and secondary dengue infections. Based on defined parameters and initial conditions, this approach allowed us to analyze the average behavior of the system.

The deterministic framework is extended to its stochastic counterpart. The stochastic differential equations (SDEs) system introduce random fluctuations or variability, capturing the inherent individual immunological variances and the probabilistic nature of the system. By considering stochasticity, we can explore how variability influences the dynamics of viremia and antibodies, offering a better understanding of the system’s behavior.

### The Deterministic framework

The deterministic framework used in our study is based on the model proposed by Sebayang, A.A et al. in [33], further analyzed by Anam, V. et al in [34]. Our study builds upon the baseline model to describe primary infections (referred to as Model A) and expands it to investigate secondary dengue infections caused by homologous (Model B) and heterologous (Model C) serotypes, considering the dynamics of viral load and antibodies, as well as the crucial role of antibody-virus complexes in infection clearance and disease enhancement. The complete modeling framework in shown in Equation 1.

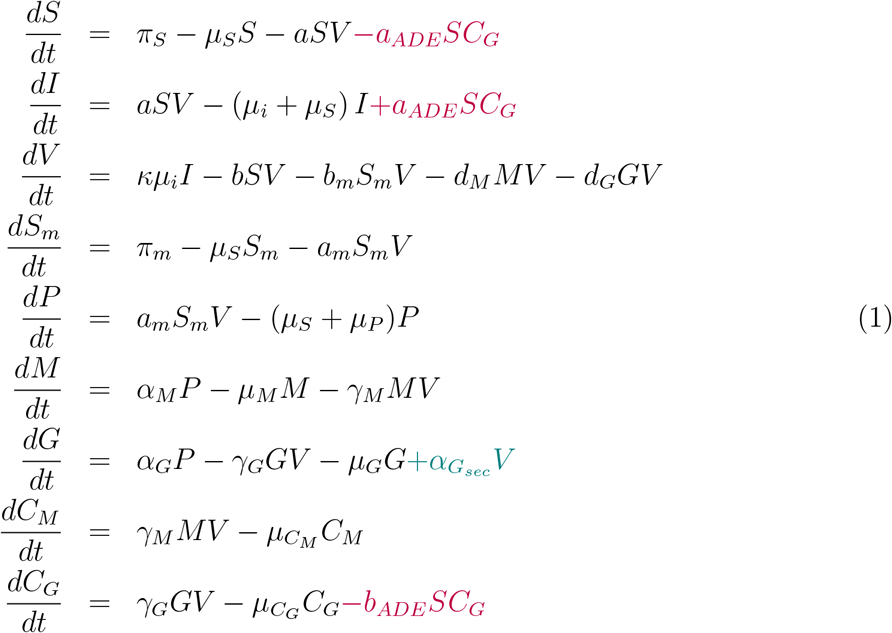

The primary infection scenario involves the following components and interactions:

1. Susceptible target cells: uninfected monocytes/dendritic cells are susceptible (*S*) to infection by encountering free virus (*V*).
2. Infected cells: When susceptible cells come into contact with the virus, they become infected cells (*I*).
3. Macrophages: uninfected macrophages become infected infected macrophages (*S*_*m*_) by encountering free virus. Upon infection, these cells act as “presenting cells” (*P*), responsible for the viral antigen presentation, triggering the production of antibodies.
4. Antibodies: Immunoglobulin M (IgM, denoted as *M*) and immunoglobulin G (IgG, denoted as *G*) are produced by the immune system upon viral antigen presentation.
5. Antibody-virus complexes: Antibodies IgM and IgG bind to the free virus, forming antibody-virus complexes. The complexes IgM-V and IgG-V are denoted as *C*_*M*_ and *C*_*G*_, respectively. These complexes are responsible for clearing the infection.

The interactions and dynamics of these components are described using a set of Ordinary Differential Equations (ODEs), which form a mathematical model to simulate the dynamics of a primary infection described by the black equations in the system 1. In the case of primary infection, the complex *C*_*M*_ is observed to lead the infection clearance, while the *C*_*G*_ is only residual.

The secondary infection scenarios are described by the same set of equations, which is extended to include a term for the pre-existing IgG antibody type (*G*_*sec*_) produced during a primary infection. The pre-existing IgG antibodies are known to be specific to a serotype, cross-reacting to the other related dengue serotypes. Dynamically, the immunological response mediated by antibodies of a secondary infection starts with the activation of these pre-existing antibodies which bind into the free virus forming the complex IgG-V (*C*_*G*_) in a much faster scale than the production of the IgM.

The model describing a homologous secondary infection includes the equations in black and the extra term in teal,

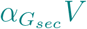

referring to the interaction of the IgG antibody and the free virus causing the new infection. In this case, the pre-existing IgG is able to neutralize the homotypic virus and clear the infection. Therefore the overall production of IgM antibodies is much lower than it is observed during a primary infection.

On the other hand, the model describing a heterologous secondary infection includes, besides the pre-existing IgG antibodies term (in teal), two extra terms shown in purple. These additional terms account for viral replication enhancement reported to occur during a secondary infection with a heterologous serotype. The term

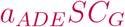

refers to the entry of *C*_*G*_ complexes into susceptible cells, generating new infected cells at rate *a*_*ADE*_. This effect is analogous to the Antibody-Dependent Enhancement ADE effect, where the pre-existing IgG antibodies are able to bind into the heterotypic virus but not to neutralize it, leading to a rapid increase of viral load in the host. The reduction of the *C*_*G*_ complexes (while generating new infected cells) during the ADE process occurs with rate *b*_*ADE*_.

### The Stochastic framework

In order to accommodate the inherent individual variances during the immunological responses to a dengue infection, we have expanded the deterministic model to its stochastic counterpart. The stochastic differential equation (SDE) serves as a diffusion approximation derived from the discrete Markov process underlying the system [46, 47].

As an illustrative example, we present the initial transition where susceptible target cells become infected upon encountering a free virus (equation 2). In this case, the state change Δ*X* is represented as Δ*X* = (*−*1, 1, 0, 0, 0, 0, 0, 0, 0), and its probability is given by:

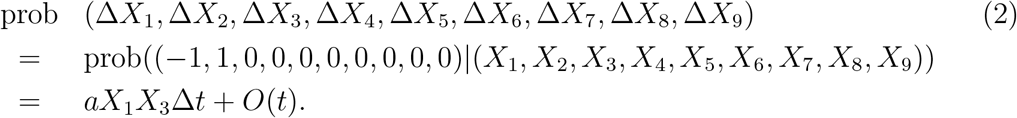

The possible state changes of the system is presented in Table S2 available in the Supplementary Material.

The construction of the stochastic differential equation model incorporates the following elements, where *dW*_*i*_ represents the weights for *i ∈* ℤ. The system SDE for model A and B is available in the Supplementary Material and the complete SDE system for model C can be written as

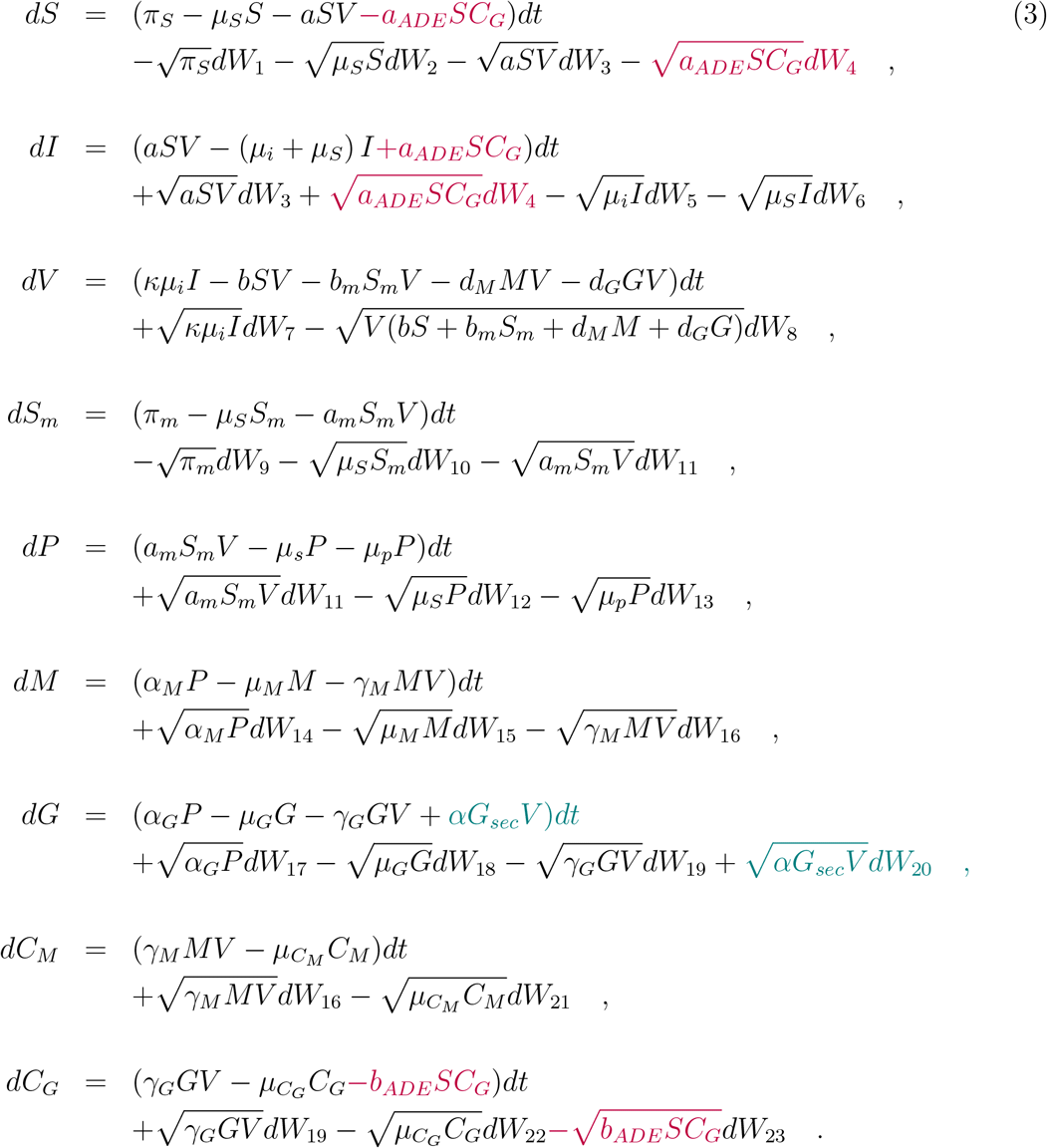

The process involved in obtaining a SDE model from a deterministic one is detailed explained in the Supplementary Material accompanying this paper.

### Modeling simulations and robustness analysis

Model simulations were conducted using MATLAB software. For the deterministic system, the simulations were obtained using the ode45 and ode15s methods, and for the stochastic simulations, the Euler-Maruyama method [47] was used. The baseline parameter values used for modeling simulations are obtained from the literature and listed in Table S1 available in the Supplementary Material.

To assess the robustness of our findings, we have compared the mean of 100 stochastic realizations with the mean field solution for each one of the models. The results obtained for Model A, Model B and Model C are shown in Figures S6-S11 available in the Supplementary Material, where a good agreement between the mean of the stochastic realizations and the mean field solutions for each model variable is observed. The baseline parameter values used for the simulations can be found in Table S1 available in the Supplementary Material.

## Conclusion

The modeling framework analyzed in this study effectively describes the immunological data for primary and secondary dengue infections. Both the deterministic and the stochastic systems are able to describe qualitatively well the immunological dynamics mediated by antibodies of primary and secondary infections.

The heterologous dengue infection dynamics account for the the antibody dependent enhancement effect, while the dynamics of homologous reinfection, which was previously thought to confer sterilizing immunity, is modeled for the first time. These homologous infections generate viremia and their potential impact in dengue transmission at the population level remains largely unexplored until now. These novel findings should be taken into consideration in future modeling research on dengue transmission and control.

Beyond its application to dengue dynamics, this modeling framework can be generalized to other infectious diseases. While adjusted to specific biological features, these models can provide accurate information of viral replication, antibody production, and infection clearance, with potential applications in decision-making, healthcare services, and industry.

## Supporting information

Supplementary Material Within-host models unravelling the dynamics of dengue reinfections

## Data Availability

The data sets analyzed in this study can be obtained from the referenced source [27] of our manuscript. Available from: https://dx.plos.org/10.1371/journal.pcbi.1004951.

## Data availability

The data sets analyzed in this study can be obtained from the referenced source [27].

## Code availability

To replicate the simulations conducted in this study, please refer to the paper for the specific modeling equations and transitions, and adapt the code accordingly using the appropriate numerical solvers and methods mentioned above. The modeling equations and transitions used in this study are available in the referenced paper. The simulations were performed using MATLAB software. For the deterministic system, the simulations were obtained using the ode45 and ode15s methods, which are standard numerical solvers for ordinary differential equations. For the stochastic simulations, the Euler-Maruyama method was employed. This method is commonly used to approximate solutions of stochastic differential equations. The code implementation for the Euler-Maruyama method can be obtained from various sources, including textbooks on numerical methods for stochastic differential equations or online repositories for MATLAB codes related to stochastic simulations.

## Competing interest

The authors declare no competing interests.

## Authors contributions

M.A, V.A and N.S conceived the study and wrote the original draft. V.A and A.K.S performed the numerical simulations. V.A., M.A and B.V.G. analyzed the data. B.V.G. and A.K.S contributed to manuscript writing and critical discussions. All authors reviewed the manuscript.

## Supplementary information

This article has accompanying supplementary file.

## Acknowledgments

This research is supported by the Basque Government through the “Mathematical Modeling Applied to Health” Project, BERC 2022-2025 program and by the Spanish Ministry of Sciences, Innovation and Universities: BCAM Severo Ochoa accreditation CEX2021-001142-S / MICIN / AEI / 10.13039/501100011033.

## Notes

### Competing Interest Statement

The authors have declared no competing interest.

### Author Declarations

The data used here were openly available to the public before the initiation of the study. The data sets analyzed in this study can be obtained from the referenced source [27] of our manuscript. Available from: https://dx.plos.org/10.1371/journal. pcbi.1004951.

## References

[1] Messina JP, Brady OJ, Golding N, Kraemer MUG, Wint GRW, Ray SE, et al. The Current and Future Global Distribution and Population at Risk of Dengue. Nature Microbiology. 2019 Jun;4(9):1508–15. Available from: https://www.nature.com/articles/s41564-019-0476-8.

[2] Waggoner JJ, Balmaseda A, Gresh L, Sahoo MK, Montoya M, Wang C, et al. Homotypic Dengue Virus Reinfections in Nicaraguan Children. Journal of Infectious Diseases. 2016 Oct;214(7):986–93. Available from: https://academic.oup.com/jid/article-lookup/doi/10.1093/infdis/jiw099.

[3] Halstead SB. Neutralization and Antibody-Dependent Enhancement of Dengue Viruses. In: Advances in Virus Research. vol. 60. Elsevier; 2003. p. 421–67. Available from: https://linkinghub.elsevier.com/retrieve/pii/S0065352703600114.

[4] Rothman AL. Cellular Immunology of Sequential Dengue Virus Infection and Its Role in Disease Pathogenesis. In: Rothman AL, editor. Dengue Virus. vol. 338. Berlin, Heidelberg: Springer Berlin Heidelberg; 2010. p. 83–98. Available from: http://link.springer.com/10.1007/978-3-642-02215-9_7.

[5] St John AL, Rathore APS. Adaptive Immune Responses to Primary and Secondary Dengue Virus Infections. Nature Reviews Immunology. 2019 Apr;19(4):218–30. Available from: https://www.nature.com/articles/s41577-019-0123-x.

[6] Guzman MG, Halstead SB, Artsob H, Buchy P, Farrar J, Gubler DJ, et al. Dengue: A Continuing Global Threat. Nature Reviews Microbiology. 2010 Dec;8(S12):S7–S16. Available from: https://www.nature.com/articles/nrmicro2460.

[7] Dejnirattisai W, Jumnainsong A, Onsirisakul N, Fitton P, Vasanawathana S, Limpitikul W, et al. Cross-Reacting Antibodies Enhance Dengue Virus Infection in Humans. Science. 2010 May;328(5979):745–8. Available from: https://www.science.org/doi/10.1126/science.1185181.

[8] Renner M, Flanagan A, Dejnirattisai W, Puttikhunt C, Kasinrerk W, Supasa P, et al. Characterization of a Potent and Highly Unusual Minimally Enhancing Antibody Directed against Dengue Virus. Nature Immunology. 2018 Nov;19(11):1248–56. Available from: https://www.nature.com/articles/s41590-018-0227-7.

[9] Hadinegoro SR, Arredondo-García JL, Capeding MR, Deseda C, Chotpitayasunondh T, Dietze R, et al. Efficacy and Long-Term Safety of a Dengue Vaccine in Regions of Endemic Disease. New England Journal of Medicine. 2015 Sep;373(13):1195–206. Available from: http://www.nejm.org/doi/10.1056/NEJMoa1506223.

[10] Biswal S, Reynales H, Saez-Llorens X, Lopez P, Borja-Tabora C, Kosalaraksa P, et al. Efficacy of a Tetravalent Dengue Vaccine in Healthy Children and Adolescents. New England Journal of Medicine. 2019 Nov;381(21):2009–19. Available from: http://www.nejm.org/doi/10.1056/NEJMoa1903869.

[11] Biswal S, Borja-Tabora C, Martinez Vargas L, Velásquez H, Theresa Alera M, Sierra V, et al. Efficacy of a Tetravalent Dengue Vaccine in Healthy Children Aged 4–16 Years: A Randomised, Placebo-Controlled, Phase 3 Trial. The Lancet. 2020 May;395(10234):1423–33. Available from: https://linkinghub.elsevier.com/retrieve/pii/S0140673620304141.

[12] Aguiar M, Stollenwerk N, Halstead SB. The Impact of the Newly Licensed Dengue Vaccine in Endemic Countries. PLOS Neglected Tropical Diseases. 2016 Dec;10(12):e0005179. Available from: https://dx.plos.org/10.1371/journal.pntd.0005179.

[13] Aguiar M, Stollenwerk N, Halstead SB. The Risks behind Dengvaxia Recommendation. The Lancet Infectious Diseases. 2016 Aug;16(8):882–3. Available from: https://linkinghub.elsevier.com/retrieve/pii/S1473309916301682.

[14] Aguiar M, Stollenwerk N. Mathematical Models of Dengue Fever Epidemiology: Multi-Strain Dynamics, Immunological Aspects Associated to Disease Severity and Vaccines. Communication in Biomathematical Sciences. 2017 Dec;1(1):1. Available from: http://journals.itb.ac.id/index.php/cbms/article/view/5767.

[15] Aguiar M. Dengue Vaccination: A More Ethical Approach Is Needed. The Lancet. 2018 May;391(10132):1769–70. Available from: https://linkinghub.elsevier.com/retrieve/pii/S0140673618308651.

[16] Aguiar M, Stollenwerk N. Dengvaxia: Age as Surrogate for Serostatus. The Lancet Infectious Diseases. 2018 Mar;18(3):245. Available from: https://linkinghub.elsevier.com/retrieve/pii/S1473309917307521.

[17] Wahala WMPB, De Silva AM. The Human Antibody Response to Dengue Virus Infection. Viruses. 2011 Nov;3(12):2374–95. Available from: http://www.mdpi.com/1999-4915/3/12/2374.

[18] Aguiar M, Stollenwerk N. The Impact of Serotype Cross-Protection on Vaccine Trials: DENVax as a Case Study. Vaccines. 2020 Nov;8(4):674. Available from: https://www.mdpi.com/2076-393X/8/4/674.

[19] Halstead SB, Katzelnick LC, Russell PK, Markoff L, Aguiar M, Dans LR, et al. Ethics of a Partially Effective Dengue Vaccine: Lessons from the Philippines. Vaccine. 2020 Jul;38(35):5572–6. Available from: https://linkinghub.elsevier.com/retrieve/pii/S0264410X20308823.

[20] Raman RS, Bhagwan Barge V, Anil Kumar D, Dandu H, Rakesh Kartha R, Bafna V, et al. A Phase II Safety and Efficacy Study on Prognosis of Moderate Pneumonia in Coronavirus Disease 2019 Patients With Regular Intravenous Immunoglobulin Therapy. The Journal of Infectious Diseases. 2021 May;223(9):1538–43. Available from: https://academic.oup.com/jid/article/223/9/1538/6135116.

[21] Rivera L, Biswal S, Sáez-Llorens X, Reynales H, López-Medina E, Borja-Tabora C, et al. Three-Year Efficacy and Safety of Takeda’s Dengue Vaccine Candidate (TAK-003). Clinical Infectious Diseases. 2022 Aug;75(1):107–17. Available from: https://academic.oup.com/cid/article/75/1/107/6381105.

[22] Aguiar M, Anam V, Blyuss KB, Estadilla CDS, Guerrero BV, Knopoff D, et al. Mathematical Models for Dengue Fever Epidemiology: A 10-Year Systematic Review. Physics of Life Reviews. 2022 Mar;40:65–92. Available from: https://linkinghub.elsevier.com/retrieve/pii/S1571064522000045.

[23] Aguiar M, Anam V, Blyuss KB, Estadilla CDS, Guerrero BV, Knopoff D, et al. Prescriptive, Descriptive or Predictive Models: What Approach Should Be Taken When Empirical Data Is Limited? Reply to Comments on “Mathematical Models for Dengue Fever Epidemiology: A 10-Year Systematic Review”. Physics of Life Reviews. 2023 Sep;46:56–64. Available from: https://linkinghub.elsevier.com/retrieve/pii/S1571064523000490.

[24] Nuraini N, Tasman H, Soewono E, Sidarto KA. A With-in Host Dengue Infection Model with Immune Response. Mathematical and Computer Modelling. 2009 Mar;49(5-6):1148–55. Available from: https://linkinghub.elsevier.com/retrieve/pii/S0895717708002732.

[25] Ben-Shachar R, Koelle K. Minimal Within-Host Dengue Models Highlight the Specific Roles of the Immune Response in Primary and Secondary Dengue Infections. Journal of The Royal Society Interface. 2015 Feb;12(103):20140886. Available from: https://royalsocietypublishing.org/doi/10.1098/rsif.2014.0886.

[26] Nikin-Beers R, Ciupe SM. The Role of Antibody in Enhancing Dengue Virus Infection. Mathematical Biosciences. 2015 May;263:83–92. Available from: https://linkinghub.elsevier.com/retrieve/pii/S0025556415000371.

[27] Clapham HE, Quyen TH, Kien DTH, Dorigatti I, Simmons CP, Ferguson NM. Modelling Virus and Antibody Dynamics during Dengue Virus Infection Suggests a Role for Antibody in Virus Clearance. PLOS Computational Biology. 2016 May;12(5):e1004951. Available from: https://dx.plos.org/10.1371/journal.pcbi.1004951.

[28] Ten Bosch QA, Singh BK, Hassan MRA, Chadee DD, Michael E. The Role of Serotype Interactions and Seasonality in Dengue Model Selection and Control: Insights from a Pattern Matching Approach. PLOS Neglected Tropical Diseases. 2016 May;10(5):e0004680. Available from: https://dx.plos.org/10.1371/journal.pntd.0004680.

[29] Ben-Shachar R, Koelle K. Transmission-Clearance Trade-Offs Indicate That Dengue Virulence Evolution Depends on Epidemiological Context. Nature Communications. 2018 Jun;9(1):2355. Available from: https://www.nature.com/articles/s41467-018-04595-w.

[30] Mapder T, Clifford S, Aaskov J, Burrage K. A Population of Bang-Bang Switches of Defective Interfering Particles Makes within-Host Dynamics of Dengue Virus Controllable. PLOS Computational Biology. 2019 Nov;15(11):e1006668. Available from: https://dx.plos.org/10.1371/journal.pcbi.1006668.

[31] Gulbudak H, Browne CJ. Infection Severity across Scales in Multi-Strain Immuno-Epidemiological Dengue Model Structured by Host Antibody Level. Journal of Mathematical Biology. 2020 May;80(6):1803–43. Available from: http://link.springer.com/10.1007/s00285-020-01480-3.

[32] Tang B, Xiao Y, Sander B, Kulkarni MA, Radam-Lac Research Team, Wu J. Modelling the Impact of Antibody-Dependent Enhancement on Disease Severity of Zika Virus and Dengue Virus Sequential and Co-Infection. Royal Society Open Science. 2020 Apr;7(4):191749. Available from: https://royalsocietypublishing.org/doi/10.1098/rsos.191749.

[33] Sebayang AA, Fahlena H, Anam V, Knopoff D, Stollenwerk N, Aguiar M, et al. Modeling Dengue Immune Responses Mediated by Antibodies: A Qualitative Study. Biology. 2021 Sep;10(9):941. Available from: https://www.mdpi.com/2079-7737/10/9/941.

[34] Anam V, Sebayang AA, Fahlena H, Knopoff D, Stollenwerk N, Soewono E, et al. Modeling Dengue Immune Responses Mediated by Antibodies: Insights on the Biological Parameters to Describe Dengue Infections. Computational and Mathematical Methods. 2022 Mar;2022:1–11. Available from: https://www.hindawi.com/journals/cmm/2022/8283239/.

[35] Nguyen NM, Thi Hue Kien D, Tuan TV, Quyen NTH, Tran CNB, Vo Thi L, et al. Host and Viral Features of Human Dengue Cases Shape the Population of Infected and Infectious Aedes Aegypti Mosquitoes. Proceedings of the National Academy of Sciences. 2013 May;110(22):9072–7. Available from: https://pnas.org/doi/full/10.1073/pnas.1303395110.

[36] Clapham HE, Tricou V, Van Vinh Chau N, Simmons CP, Ferguson NM. Within-Host Viral Dynamics of Dengue Serotype 1 Infection. Journal of The Royal Society Interface. 2014 Jul;11(96):20140094. Available from: https://royalsocietypublishing.org/doi/10.1098/rsif.2014.0094.

[37] Nguyen NM, Tran CNB, Phung LK, Duong KTH, Huynh HLA, Farrar J, et al. A Randomized, Double-Blind Placebo Controlled Trial of Balapiravir, a Polymerase Inhibitor, in Adult Dengue Patients. The Journal of Infectious Diseases. 2013 May;207(9):1442–50. Available from: https://academic.oup.com/jid/article-lookup/doi/10.1093/infdis/jis470.

[38] Vaughn DW, Green S, Kalayanarooj S, Innis BL, Nimmannitya S, Suntayakorn S, et al. Dengue Viremia Titer, Antibody Response Pattern, and Virus Serotype Correlate with Disease Severity. The Journal of Infectious Diseases. 2000 Jan;181(1):2–9. Available from: https://academic.oup.com/jid/article-lookup/doi/10.1086/315215.

[39] Martial J, Dussart P, Plumelle Y, Moravie V, Verlaeten O, Najioullah F, et al. Influence of the Dengue Serotype, Previous Dengue Infection, and Plasma Viral Load on Clinical Presentation and Outcome During a Dengue-2 and Dengue-4 Co-Epidemic. The American Journal of Tropical Medicine and Hygiene. 2008 Jun;78(6):990–8. Available from: https://ajtmh.org/doi/10.4269/ajtmh.2008.78.990.

[40] Wang WK, Chao DY, Kao CL, Wu HC, Liu YC, Li CM, et al. High Levels of Plasma Dengue Viral Load during Defervescence in Patients with Dengue Hemorrhagic Fever: Implications for Pathogenesis. Virology. 2003 Jan;305(2):330–8. Available from: https://linkinghub.elsevier.com/retrieve/pii/S0042682202917046.

[41] Guilarde AO, Turchi MD, Jr JBS, Feres VCR, Rocha B, Levi JE, et al. Dengue and Dengue Hemorrhagic Fever among Adults: Clinical Outcomes Related to Viremia, Serotypes, and Antibody Response. The Journal of Infectious Diseases. 2008 Mar;197(6):817–24. Available from: https://academic.oup.com/jid/article-lookup/doi/10.1086/528805.

[42] Boonnak K, Dambach KM, Donofrio GC, Tassaneetrithep B, Marovich MA. Cell Type Specificity and Host Genetic Polymorphisms Influence Antibody-Dependent Enhancement of Dengue Virus Infection. Journal of Virology. 2011 Feb;85(4):1671–83. Available from: https://journals.asm.org/doi/10.1128/JVI.00220-10.

[43] Bournazos S, Gupta A, Ravetch JV. The Role of IgG Fc Receptors in Antibody-Dependent Enhancement. Nature Reviews Immunology. 2020 Oct;20(10):633–43. Available from: https://www.nature.com/articles/s41577-020-00410-0.

[44] Guzman MG, Harris E. Dengue. The Lancet. 2015 Jan;385(9966):453–65. Available from: https://linkinghub.elsevier.com/retrieve/pii/S0140673614605729.

[45] Changal KH, Raina AH, Raina A, Raina M, Bashir R, Latief M, et al. Differentiating Secondary from Primary Dengue Using IgG to IgM Ratio in Early Dengue: An Observational Hospital Based Clinico-Serological Study from North India. BMC Infectious Diseases. 2016 Dec;16(1):715. Available from: http://bmcinfectdis.biomedcentral.com/articles/10.1186/s12879-016-2053-6.

[46] Allen EJ, Allen LJS, Arciniega A, Greenwood PE. Construction of Equivalent Stochastic Differential Equation Models. Stochastic Analysis and Applications. 2008 Mar;26(2):274–97. Available from: http://www.tandfonline.com/doi/abs/10.1080/07362990701857129.

[47] Kloeden PE, Platen E. Numerical Solution of Stochastic Differential Equations. Berlin, Heidelberg: Springer Berlin Heidelberg; 1992. Available from: http://link.springer.com/10.1007/978-3-662-12616-5.

