## Supplementary Material Within-host models unravelling the dynamics of dengue reinfections for "Within-host models unravelling the dynamics of dengue reinfections"

#### 1 Data classification deviating from prior studies

Our analysis of the data from the study conducted by Nguyen et al. [1] has revealed discrepancies in the classifications used in previous modeling studies, raising uncertainties and highlighting the importance of careful data interpretation.

To address the uncertainties in data classification, we have investigated these cases using our models. The results are shown in the following figures, supporting the current classification of these samples. The baseline parameters used for modeling simulations are listed in Table S1.

Upon data inspection, we reclassified five samples as follows.

#### 1.1 From DENV1 infections

Patient ID 225: previously treated as secondary infection, this subject was reclassified as primary dengue infection. The sample presents high viral titers, high IgM antibody concentrations, and low IgG antibody concentrations. The IgG concentration levels are observed to be near the threshold point, eventually resulting in unspecific serological results. This sample was tested with Model A, Model B and Model C, as shown in Figure S1.

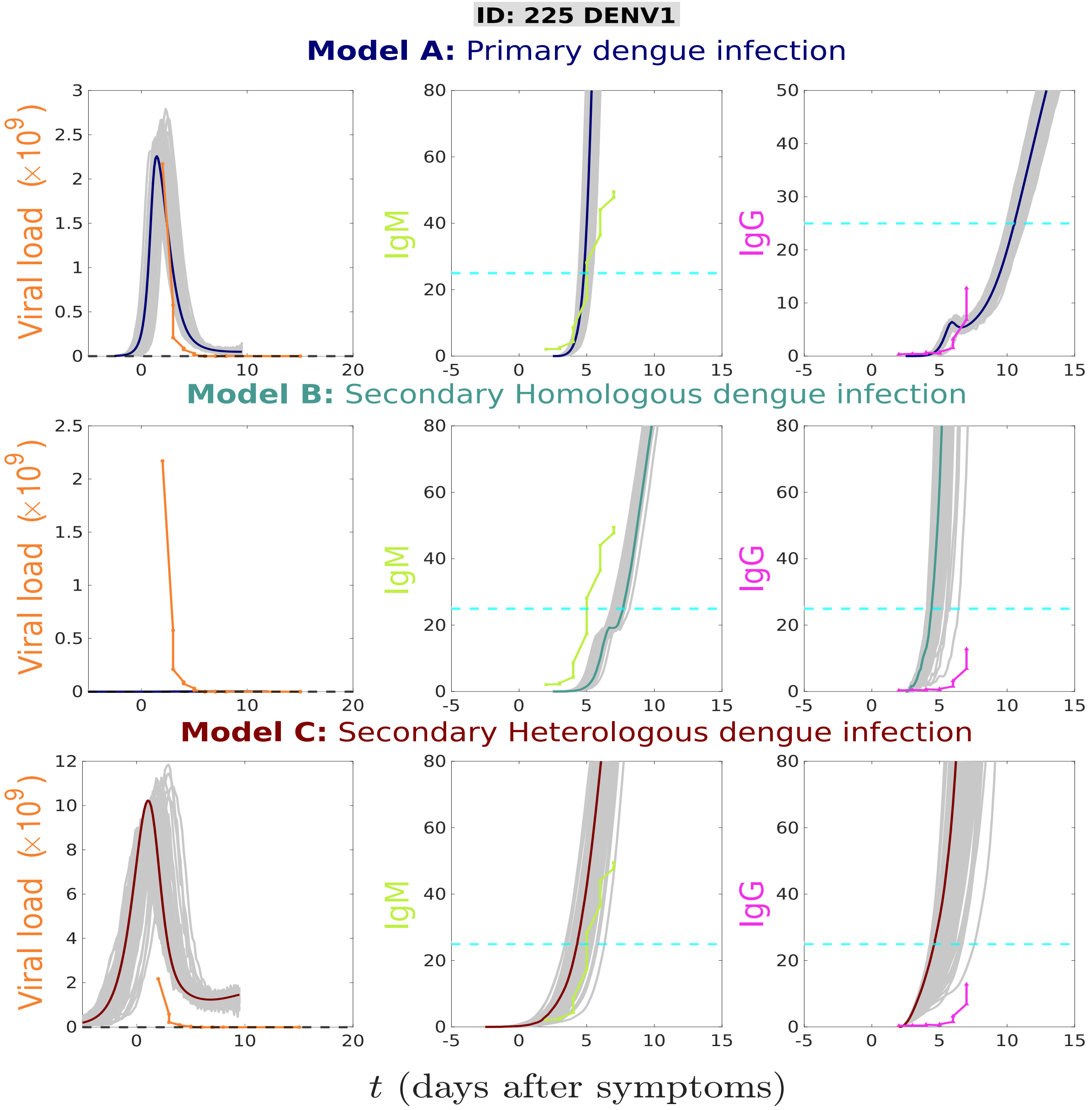


Figure S1: Stochastic realizations (grey lines) and the deterministic solutions (full lines) for models A, B, and C are compared for patient ID 225. Viral load is shown in orange, whereas antibody concentrations are shown in green (IgM) and in magenta (IgG). Model A describe well the dynamics of all variables. While Model B shows low stochastic realizations for viral load (close to zero), Model C shows very high stochastic realizations as compared to the data.

Patient ID 208: previously excluded from the analysis conducted in [2], this subject has been classified as a secondary homologous infection. The sample presents viral titer well below the detection threshold, along with high IgG antibody concentrations levels. This sample was tested with Model B and Model C, as shown in Figure S2.

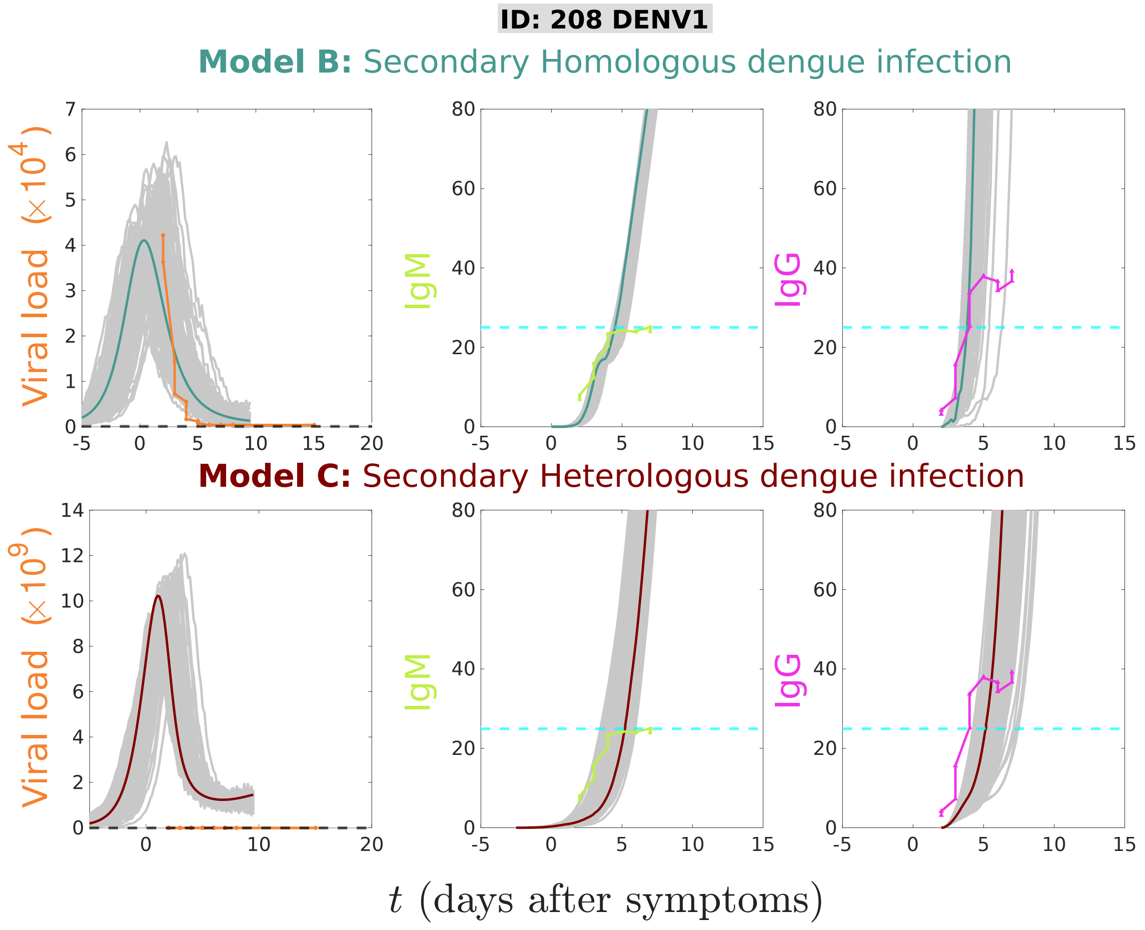


Figure S2: Stochastic realizations (grey lines) and the deterministic solutions (full lines) for models A, B, and C are compared for patient ID 208. Viral load is shown in orange, whereas antibody concentrations are shown in green (IgM) and in magenta (IgG). Model B describe well the dynamics of all variables, while Model C shows very high stochastic realizations as compared to the data.

#### 1.2 From DENV2 infection

Patient ID 106: previously treated a secondary heterologous infection, this subject has been reclassified as secondary homologous infection. The sample presents viral titer well below the detection threshold, along with high IgG antibody concentrations levels. This sample was tested with Model B and Model C, as shown in Figure S3.

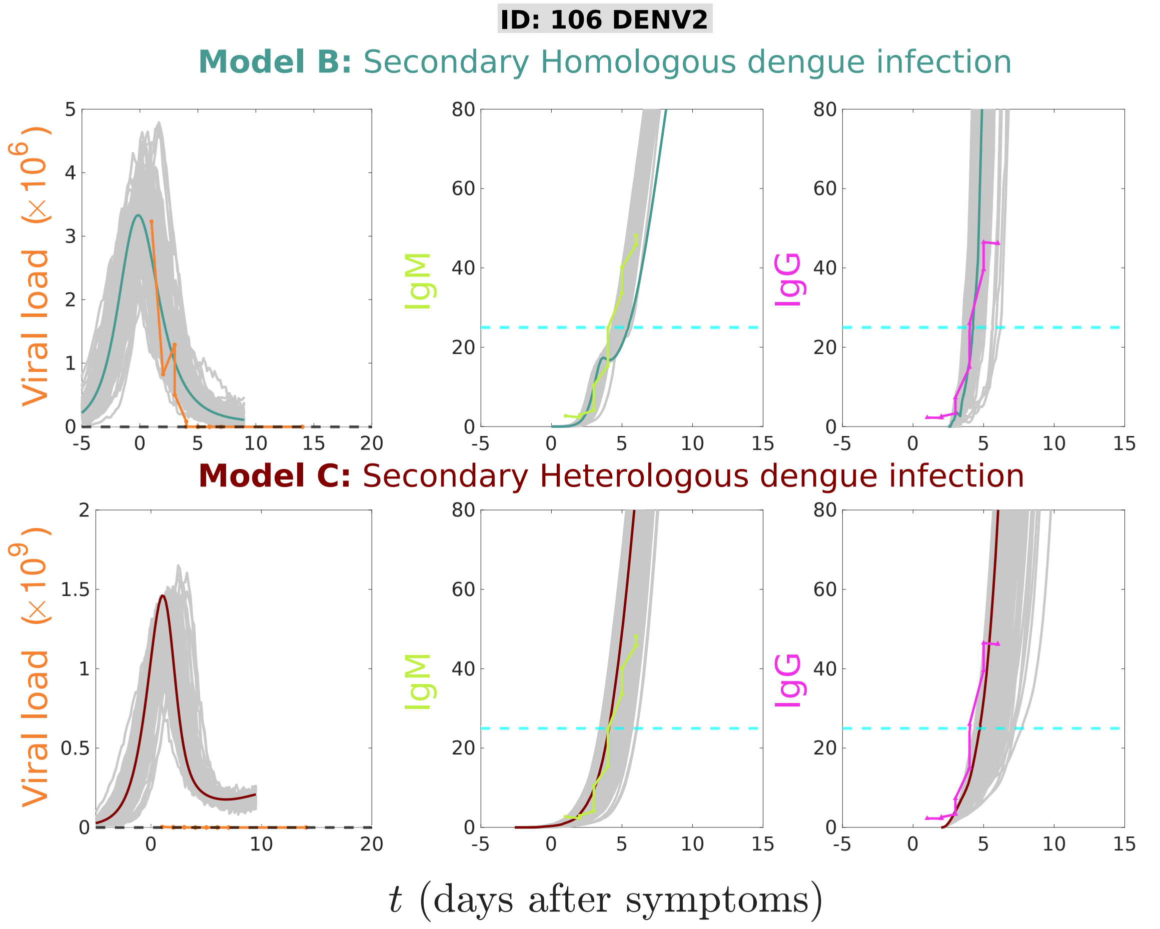


Figure S3: Stochastic realizations (grey lines) and the deterministic solutions (full lines) for models A, B, and C are compared for patient ID 106. Viral load is shown in orange, whereas antibody concentrations are shown in green (IgM) and in magenta (IgG). Model B describe well the dynamics of all variables, while Model C shows very high stochastic realizations as compared to the data.

#### 1.3 From DENV4 infections

Patient ID 108: previously treated as secondary infection, this subject has been reclassified as primary infection. This sample presents high IgM levels with the concentration of IgG antibodies near the threshold of detection, with possible unspecific serological result. This sample was tested with Model A, Model B and Model C, as shown in Figure S4

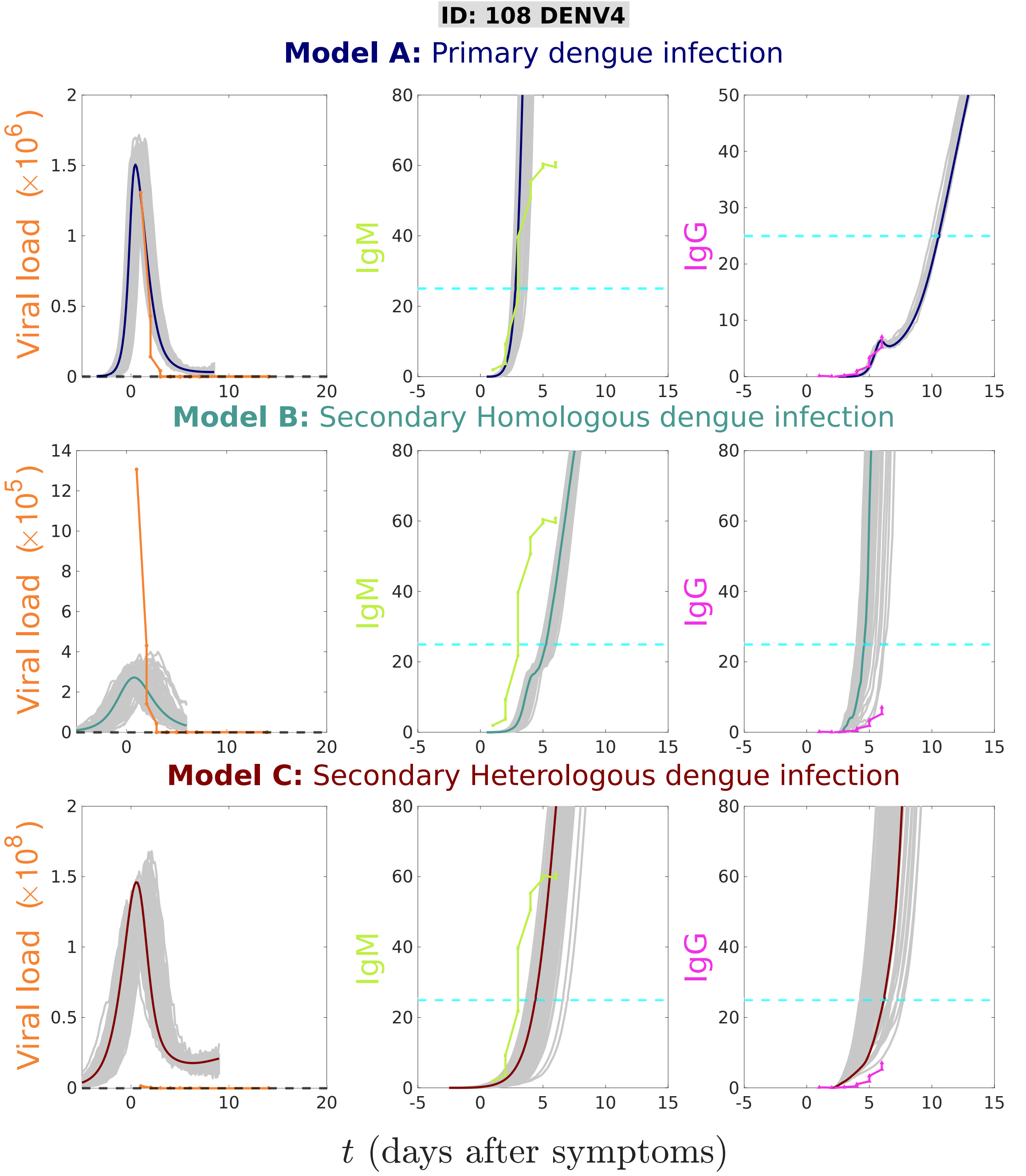


Figure S4: Stochastic realizations (grey lines) and the deterministic solutions (full lines) for models A, B, and C are compared for patient ID 108. Viral load is shown in orange, whereas antibody concentrations are shown in green (IgM) and in magenta (IgG). Model A describe well the dynamics of all variables. While Model B shows low stochastic realizations for viral load (close to zero), Model C shows very high stochastic realizations as compared to the data.

Patient ID 110: previously treated a secondary heterologous infection, this subject has been reclassified as secondary homologous infection. The basis for this classification is the observation of low viremia and high IgG and IgM concentration levels in the patient's sample. This sample was tested with Model B and Model C, as shown in Figure S5.

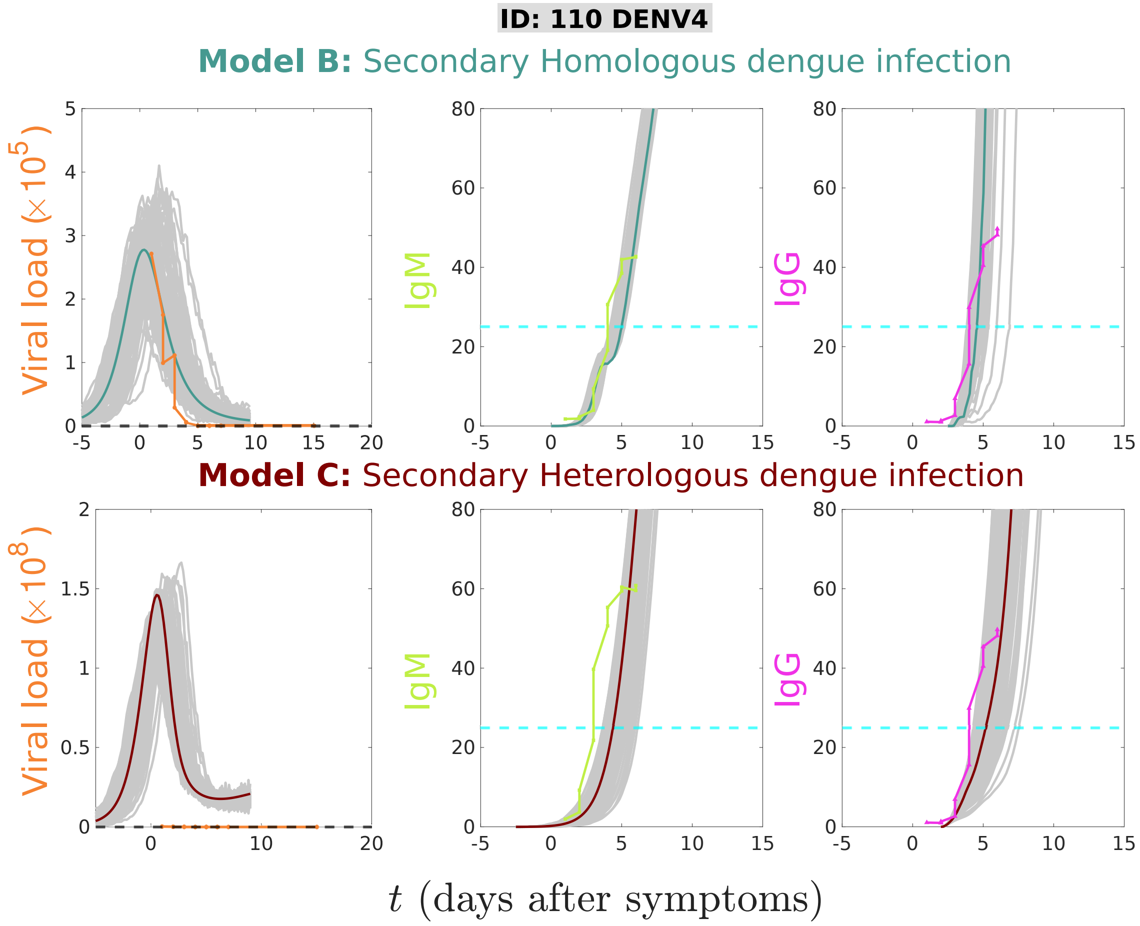


Figure S5: Stochastic realizations (grey lines) and the deterministic solutions (full lines) for models A, B, and C are compared for patient ID 110. Viral load is shown in orange, whereas antibody concentrations are shown in green (IgM) and in magenta (IgG). Model B describe well the dynamics of all variables, while Model C shows very high stochastic realizations as compared to the data.

Table S1: Baseline parameter values used for the modeling simulations

| **Parameters** | **Values** | **Description** | **Reference** |
|---|---|---|---|
| $\pi_S$ | 600 | production rate of susceptible target cells (monocytes/dendritic cells) per day | [3, 4] |
| $\pi_M$ | 300 | production rate of susceptible macrophages per day | [3, 4] |
| $\mu_S$ | 1/30 | natural mortality of susceptible target cells (monocytes/dendritic cells) per day | [5] |
| $\mu_i$ | 2 | decay rate of infected cells (monocytes/dendritic cells) per day | [4] |
| $\mu_P$ | $\mu_i/10$ | decay rate of presenting cells (infected macrophages) per day | [4] |
| $a$ | 0.2 | infection rate of susceptible target cells per day | [4] |
| $a_m$ | 0.2 | infection rate of susceptible macrophages per day | [4] |
| $b$ | $12a$ | removal rate of viral particles infecting susceptible target cells per day | [4] |
| $b_m$ | $12a$ | removal rate of viral particles infecting susceptible macrophages per day | [4] |
| $\kappa$ | 50 | viral replication factor | [6] |
| $\alpha_M$ | 10 | production rate of IgM antibody per day | [4] |
| $\alpha_G$ | 1.5 | production rate of IgG antibody per day | [4] |
| $\alpha_{G_{sec}}$ | $(2000)\alpha_G$ | production rate of pre-existing IgG antibody per day | [4] |
| $\gamma_M = \gamma_G$ | 0.06 | binding rate of antibodies (IgM and IgG) per day | [4] |
| $d_M$ | $4 \cdot \gamma_M$ | average number of free virus particle forming the virus-IgM complex | [4] |
| $d_G$ | $\gamma_G$ | average number of free virus particle forming the virus-IgG complex | [4] |
| $\mu_M$ | 0.03 | decay rate of antibody IgM per day | [7] |
| $\mu_G$ | 1/365 | decay rate of antibody IgG per day | [7] |
| $\mu_{CM} = \mu_{CG}$ | 1 | decay rate of virus-antibody complexes $C_M$ and $C_G$ per day | [4] |
| $S(t_0)$ | $\pi_S/\mu_S$ | initial value for susceptible target cells (monocytes and dendritic) | [5] |
| $S_m(t_0)$ | $\pi_M/\mu_S$ | initial value for susceptible macrophages | [5] |
| $V(t_0)$ | 3 | initial value of free viral particles upon infection | [4] |

### 2 Methods

In order to account for the inherent individual immunological variances during the infection process, the deterministic framework is extended to its stochastic counterpart. The stochastic differential equation (SDE) serves as a diffusion approximation derived from the discrete Markov process underlying the system [8, 9].

The SDE is a differential equation that describes the evolution of a system over time, considering both deterministic and probabilistic components.

The general form of an SDE is given by:

$$dX(t) = f(X(t), t)dt + g(X(t), t)dW(t)$$

where:

$X(t)$ is the state variable of the system at time $t$.

$f(X(t), t)$ represents the deterministic part of the model, describing how the state variable changes over time in a deterministic manner.

$g(X(t), t)$ represents the state dependence of the noise, determining the amplitude of the noise on the system.

$dW(t)$ is the differential of a Wiener process (Brownian motion) and represents the random noise in the system.

The transitions for state changes are defined in Table S2.

Table S2: Transitions for models A, B and C.

| state transitions | biological mechanisms |
|---|---|
| $\mathrm{S} \xrightarrow{\pi_{\mathrm{S}}} \mathrm{S}\uparrow$ | constant input of susceptible target cells |
| $\mathrm{S} \xrightarrow{\mu_{\mathrm{S}}} \mathrm{S}\downarrow$ | natural mortality of susceptible target cells |
| $\mathrm{S}+\mathrm{V} \xrightarrow{a} \mathrm{S}\downarrow + \mathrm{I}\uparrow$ | susceptible cells meeting free virus particles becoming infected cells |
| $\mathrm{I} \xrightarrow{\mu_i} \mathrm{I}\downarrow$ | infected target cells mortality |
| $\mathrm{I} \xrightarrow{\mu_{\mathrm{S}}} \mathrm{I}\downarrow$ | natural mortality of infected macrophages |
| $\mathrm{V} \xrightarrow{\kappa} \mathrm{V}\uparrow$ | viral particles released when infected target cells die |
| $\mathrm{S}+\mathrm{S}_m+\mathrm{V}+\mathrm{M}+\mathrm{G} \xrightarrow{\mathrm{b}+\mathrm{b}_m+\mathrm{d}_{\mathrm{M}}+\mathrm{d}_{\mathrm{G}}} \mathrm{V}\downarrow$ | removal rate of virus particles |
| $\mathrm{S}_m \xrightarrow{\pi_m} \mathrm{S}_m\uparrow$ | constant input of susceptible macrophages |
| $\mathrm{S}_m \xrightarrow{\mu_{\mathrm{S}}} \mathrm{S}_m\downarrow$ | natural mortality of susceptible macrophages |
| $\mathrm{S}_m+\mathrm{V} \xrightarrow{a_m} \mathrm{S}_m\downarrow + \mathrm{P}\uparrow$ | infected macrophages becoming presenting cells |
| $\mathrm{P} \xrightarrow{\mu_{\mathrm{S}}} \mathrm{P}\downarrow$ | natural mortality of infected macrophages |
| $\mathrm{P} \xrightarrow{\mu_{\mathrm{P}}} \mathrm{P}\downarrow$ | infected presenting cells mortality |
| $\mathrm{M} \xrightarrow{\alpha_{\mathrm{M}}} \mathrm{M}\uparrow$ | production of antibody IgM |
| $\mathrm{M} \xrightarrow{\mu_{\mathrm{M}}} \mathrm{M}\downarrow$ | decay of antibody IgM |
| $\mathrm{M}+\mathrm{V} \xrightarrow{\gamma_{\mathrm{M}}} \mathrm{C}_{\mathrm{M}}\uparrow + \mathrm{M}\downarrow$ | production of IgM-V complexes |
| $\mathrm{G} \xrightarrow{\alpha_{\mathrm{G}}} \mathrm{G}\uparrow$ | production of antibody IgG |
| $\mathrm{G} \xrightarrow{\mu_{\mathrm{G}}} \mathrm{G}\downarrow$ | decay of antibody IgG |
| $\mathrm{G}+\mathrm{V} \xrightarrow{\gamma_{\mathrm{G}}} \mathrm{C}_{\mathrm{G}}\uparrow + \mathrm{G}\downarrow$ | production of IgG-V complexes |
| $\mathrm{C}_{\mathrm{M}} \xrightarrow{\mu_{\mathrm{CM}}} \mathrm{C}_{\mathrm{M}}\downarrow$ | decay of IgM-V complexes |
| $\mathrm{C}_{\mathrm{G}} \xrightarrow{\mu_{\mathrm{CG}}} \mathrm{C}_{\mathrm{G}}\downarrow$ | decay of IgG-V complexes |
| $\mathrm{G}+\mathrm{V} \xrightarrow{\alpha_{\mathrm{Gsec}}} \mathrm{G}\uparrow$ | production of pre-existing serotype-specific IgG antibodies |
| $\mathrm{S}+\mathrm{C}_{\mathrm{G}} \xrightarrow{a_{ADE}} \mathrm{I}\uparrow + \mathrm{S}\downarrow$ | pre-existing IgG-V complexes infecting susceptible target cells (ADE process) |
| $\mathrm{S}+\mathrm{C}_{\mathrm{G}} \xrightarrow{\mathrm{b}_{\mathrm{ADE}}} \mathrm{V}\downarrow$ | removal of pre-existing IgG-V complexes |

#### 2.1 Construction of the Stochastic Differential Equation (SDE) system

##### 2.1.1 Model A

Following the method developed by Yuan et al. [10], the deterministic Model A (Equation System 6) is extended to its stochastic counterpart.

Let

$$X(t) = (X_1(t), X_2(t), X_3(t), X_4(t), X_5(t), X_6(t), X_7(t), X_(8), X_9(t))^T$$

be a continuous random variable for

$$[S(t), I(t), V(t), S_m(t), P(t), M(t), G(t), C_M(t), C_G(t)]^T,$$

where $T$ denotes transpose of the matrix.

Further, let $\Delta X = X(t+\Delta t) - X(t) = (\Delta X_1, \Delta X_2, \Delta X_3, \Delta X_4 \ldots, \Delta X_9.)^T$ denotes the random vector for the change in random variables during time interval $\Delta t$.

The construction of a stochastic process is illustrated by considering the scenario where susceptible target cells $S$ encounter free virus $V$, becoming an infected cell $I$, as shown in equation $\frac{dS}{dt}$ of Equation System 1.

$$\begin{aligned}
\frac{dS}{dt} &= \pi_S - \mu_S S - aSV \\
\frac{dI}{dt} &= aSV - (\mu_i + \mu_S)\, I \\
\frac{dV}{dt} &= \kappa\mu_i I - bSV - b_m S_m V - d_M MV - d_G GV \\
\frac{dS_m}{dt} &= \pi_m - \mu_S S_m - a_m S_m V \\
\frac{dP}{dt} &= a_m S_m V - (\mu_S + \mu_P)P \\
\frac{dM}{dt} &= \alpha_M P - \mu_M M - \gamma_M MV \\
\frac{dG}{dt} &= \alpha_G P - \gamma_G GV - \mu_G G \\
\frac{dC_M}{dt} &= \gamma_M MV - \mu_{C_M} C_M \\
\frac{dC_G}{dt} &= \gamma_G GV - \mu_{C_G} C_G
\end{aligned} \tag{1}$$

For this specific case, the state change $\Delta X$ is denoted by $\Delta X = (-1, 1, 0, 0, 0, 0, 0, 0, 0)$ and its probability is given by

$$\begin{aligned}
&\text{prob} \quad (\Delta X_1, \Delta X_2, \Delta X_3, \Delta X_4, \Delta X_5, \Delta X_6, \Delta X_7, \Delta X_8, \Delta X_9) \qquad (2)\\
&= \quad \text{prob}((-1, 1, 0, 0, 0, 0, 0, 0, 0, 0)|(X_1, X_2, X_3, X_4, X_5, X_6, X_7, X_8, X_9)) \\
&= \quad aX_1X_3\Delta t + O(t).
\end{aligned}$$

The state changes and their probabilities are presented in Table S3.

Table S3: Model A: changes of states and their probabilities.

| Possible state change | Probability of state change |
|---|---|
| $(\Delta X)_1 = (1,0,0,0,0,0,0,0,0)^T$ | $P_1 = \pi_S \Delta t + O(\Delta t)$ |
| $(\Delta X)_2 = (-1,0,0,0,0,0,0,0,0)^T$ | $P_2 = \mu_S X_1 \Delta t + O(\Delta t)$ |
| $(\Delta X)_3 = (-1,1,0,0,0,0,0,0,0)^T$ | $P_3 = a X_1 X_3 \Delta t + O(\Delta t)$ |
| $(\Delta X)_4 = (0,-1,0,0,0,0,0,0,0)^T$ | $P_4 = \mu_i X_2 \Delta t + O(\Delta t)$ |
| $(\Delta X)_5 = (0,-1,0,0,0,0,0,0,0)^T$ | $P_5 = \mu_S X_2 \Delta t + O(\Delta t)$ |
| $(\Delta X)_6 = (1,0,0,0,0,0,0,0,0)^T$ | $P_6 = \kappa \mu_i X_2 \Delta t + O(\Delta t)$ |
| $(\Delta X)_7 = (0,0,-1,0,0,0,0,0,0)^T$ | $P_7 = X_3(bX_1 + b_m X_4 + d_M X_6 + d_G X_7)\Delta t + O(\Delta t)$ |
| $(\Delta X)_8 = (0,0,0,1,0,0,0,0,0)^T$ | $P_8 = \pi_m \Delta t + O(\Delta t)$ |
| $(\Delta X)_9 = (0,0,0,-1,0,0,0,0,0)^T$ | $P_9 = \mu_S X_4 \Delta t + O(\Delta t)$ |
| $(\Delta X)_{10} = (0,0,0,-1,1,0,0,0,0)^T$ | $P_{10} = a_m X_3 X_4 \Delta t + O(\Delta t)$ |
| $(\Delta X)_{11} = (0,0,0,0,-1,0,0,0,0)^T$ | $P_{11} = \mu_S X_5 \Delta t + O(\Delta t)$ |
| $(\Delta X)_{12} = (0,-1,0,0,0,0,0,0,0)^T$ | $P_{12} = \mu_p X_5 \Delta t + O(\Delta t)$ |
| $(\Delta X)_{13} = (0,0,0,0,0,1,0,0,0)^T$ | $P_{13} = \alpha_M X_5 \Delta t + O(\Delta t)$ |
| $(\Delta X)_{14} = (0,0,0,0,0,-1,0,0,0)^T$ | $P_{14} = \mu_M X_6 \Delta t + O(\Delta t)$ |
| $(\Delta X)_{15} = (0,0,0,0,0,-1,0,1,0)^T$ | $P_{15} = \gamma_M X_3 X_6 \Delta t + O(\Delta t)$ |
| $(\Delta X)_{16} = (0,0,0,0,0,0,1,0,0)^T$ | $P_{16} = \alpha_G X_5 \Delta t + O(\Delta t)$ |
| $(\Delta X)_{17} = (0,0,0,0,0,0,-1,0,0)^T$ | $P_{17} = \mu_G X_7 \Delta t + O(\Delta t)$ |
| $(\Delta X)_{18} = (0,0,0,0,0,0,-1,0,1)^T$ | $P_{18} = \gamma_G X_3 X_7 \Delta t + O(\Delta t)$ |
| $(\Delta X)_{19} = (0,0,0,0,0,0,0,-1,0)^T$ | $P_{19} = \mu_{CM} X_8 \Delta t + O(\Delta t)$ |
| $(\Delta X)_{20} = (0,0,0,0,0,0,0,0,-1)^T$ | $P_{20} = \mu_{CG} X_9 \Delta t + O(\Delta t)$ |
| $(\Delta X)_{21} = (0,0,0,0,0,0,0,0,0)^T$ | $P_{21} = 1 - \sum_{i=1}^{20} \Delta t + O(\Delta t)$ |

By neglecting the terms equal and higher to $O^2(\Delta t)$, the expectation change $E(\Delta X)$ and its covariance matrix $R(\Delta X)$ associated with $\Delta X$ is given by

$$
\begin{aligned}
E(\Delta X) &= \sum_{i=1}^{20} P_i (\Delta X)_i \Delta t \\
&= \begin{pmatrix}
\pi_S - \mu_S X_1 - a X_1 X_3 \\
a X_1 X_3 - \mu_i X_2 - \mu_S X_2 \\
\kappa \mu_i X_2 - X_3(bX_1 + b_m X_4 + d_M X_6 + d_G X_7) \\
\pi_m - \mu_S X_4 - a_m X_3 X_4 \\
a_m X_3 X_4 - \mu_s X_5 - \mu_p X_5 \\
\alpha_M X_5 - \mu_M X_6 - \gamma_M X_3 X_6 \\
\alpha_G X_5 - \mu_G X_7 - \gamma_G X_3 X_7 \\
\gamma_M X_3 X_6 - \mu_{CM} X_8 \\
\gamma_G X_3 X_7 - \mu_{CG} X_9
\end{pmatrix}.
\end{aligned}
$$

Since the covariance matrix is $R(\Delta X) = E((\Delta X)(\Delta X)^T) - E(\Delta X)E((\Delta X)^T)$ and $E(\Delta X)E((\Delta X)^T) = f(X)(f(X)^T)$, it can be approximated with diffusion matrix $\Omega$

times $\Delta t$ by neglecting the term of $(\Delta t)^2$ such that

$$
\begin{aligned}
E((\Delta X)(\Delta X)^T) &= \sum_{i=1}^{20} P_i((\Delta X)_i(\Delta X)_i^T)\Delta t \\
&= \begin{pmatrix}
R_{11} & R_{12} & 0 & 0 & 0 & 0 & 0 & 0 & 0 \\
R_{21} & R_{22} & 0 & 0 & 0 & 0 & 0 & 0 & 0 \\
0 & 0 & R_{33} & 0 & 0 & 0 & 0 & 0 & 0 \\
0 & 0 & 0 & R_{44} & R_{45} & 0 & 0 & 0 & 0 \\
0 & 0 & 0 & R_{54} & R_{55} & 0 & 0 & 0 & 0 \\
0 & 0 & 0 & 0 & 0 & R_{66} & 0 & R_{68} & 0 \\
0 & 0 & 0 & 0 & 0 & 0 & R_{77} & 0 & R_{79} \\
0 & 0 & 0 & 0 & 0 & R_{86} & 0 & R_{88} & 0 \\
0 & 0 & 0 & 0 & 0 & 0 & R_{97} & 0 & R_{99}
\end{pmatrix} \cdot \Delta t \\
&= \Omega \cdot \Delta t,
\end{aligned}
$$

where the above diffusion matrix is symmetric, positive-definite and each component of this $9 \times 9$ diffusion matrix are given by

$$
\begin{aligned}
R_{11} &= P_1 + P_2 + P_3 = \pi_S - \mu_S X_1 + aX_1X_3, \\
R_{12} &= R_{21} = -P_3 = -aX_1X_3, \\
R_{22} &= P_3 + P_4 + P_5 = aX_1X_3 + \mu_i X_2 + \mu_S X_2, \\
R_{33} &= P_6 + P_7 = \kappa\mu_i X_2 + X_3(bX_1 + b_m X_4 + d_M X_6 + d_G X_7), \\
R_{44} &= P_8 + P_9 + P_{10} = \pi_m - \mu_S X_4 + a_m X_3 X_4, \\
R_{45} &= R_{54} = -P_{10} = -a_m X_3 X_4, \\
R_{55} &= P_{10} + P_{11} + P_{12} = a_m X_3 X_4 + \mu_S X_5 + \mu_p X_5, \\
R_{66} &= P_{13} + P_{14} + P_{15} = \alpha_M X_5 + \mu_M X_6 + \gamma_M X_3 X_6, \\
R_{68} &= R_{86} = -P_{15} = -\gamma_M X_3 X_6, \\
R_{77} &= P_{16} + P_{17} + P_{18} = \alpha_G X_5 + \mu_G X_7 + \gamma_G X_3 X_7, \\
R_{79} &= R_{97} = -P_{18} = -\gamma_G X_3 X_7, \\
R_{88} &= P_{15} + P_{19} = \gamma_M X_3 X_6 + \mu_{CM} X_8, \\
R_{99} &= P_4 + P_6 = \gamma_G X_3 X_7 + \mu_{CG} X_9.
\end{aligned}
$$

where

$$
\begin{aligned}
&U_{11} = \sqrt{P_1},\ U_{12} = -\sqrt{P_2},\ U_{13} = -\sqrt{P_3},\ U_{23} = \sqrt{P_3},\ U_{24} = -\sqrt{P_4},\ U_{25} = -\sqrt{P_5}, \\
&U_{36} = \sqrt{P_6},\ U_{37} = -\sqrt{P_7},\ U_{48} = \sqrt{P_8},\ U_{49} = -\sqrt{P_9},\ U_{4,10} = -\sqrt{P_{10}},\ U_{5,10} = \sqrt{P_{10}}, \\
&U_{5,11} = -\sqrt{P_{11}},\ U_{5,12} = -\sqrt{P_{12}},\ U_{6,13} = \sqrt{P_{13}},\ U_{6,14} = -\sqrt{P_{14}},\ U_{6,15} = -\sqrt{P_{15}}, \\
&U_{8,15} = \sqrt{P_{15}},\ U_{7,16} = \sqrt{P_{16}},\ U_{7,17} = -\sqrt{P_{17}},\ U_{7,18} = -\sqrt{P_{18}}, \\
&U_{9,18} = \sqrt{P_{18}},\ U_{8,19} = -\sqrt{P_{19}}, \\
&U_{9,20} = -\sqrt{P_{20}}.
\end{aligned}
$$

$$U = \begin{bmatrix}
U_{11} & U_{12} & U_{13} & 0 & 0 & 0 & 0 & 0 & 0 & 0 & 0 & 0 & 0 & 0 & 0 & 0 & 0 & 0 & 0 & 0 \\
0 & 0 & U_{23} & U_{24} & U_{25} & 0 & 0 & 0 & 0 & 0 & 0 & 0 & 0 & 0 & 0 & 0 & 0 & 0 & 0 & 0 \\
0 & 0 & 0 & 0 & 0 & U_{36} & U_{37} & 0 & 0 & 0 & 0 & 0 & 0 & 0 & 0 & 0 & 0 & 0 & 0 & 0 \\
0 & 0 & 0 & 0 & 0 & 0 & 0 & U_{48} & U_{49} & U_{4,10} & 0 & 0 & 0 & 0 & 0 & 0 & 0 & 0 & 0 & 0 \\
0 & 0 & 0 & 0 & 0 & 0 & 0 & 0 & 0 & U_{5,10} & U_{5,11} & U_{5,12} & 0 & 0 & 0 & 0 & 0 & 0 & 0 & 0 \\
0 & 0 & 0 & 0 & 0 & 0 & 0 & 0 & 0 & 0 & 0 & 0 & U_{6,13} & U_{6,14} & U_{6,15} & 0 & 0 & 0 & 0 & 0 \\
0 & 0 & 0 & 0 & 0 & 0 & 0 & 0 & 0 & 0 & 0 & 0 & 0 & 0 & 0 & U_{7,16} & U_{7,17} & U_{7,18} & 0 & 0 \\
0 & 0 & 0 & 0 & 0 & 0 & 0 & 0 & 0 & 0 & 0 & 0 & 0 & 0 & U_{8,15} & 0 & 0 & 0 & U_{8,19} & 0 \\
0 & 0 & 0 & 0 & 0 & 0 & 0 & 0 & 0 & 0 & 0 & 0 & 0 & 0 & 0 & 0 & 0 & U_{9,18} & 0 & U_{9,20}
\end{bmatrix}$$

Following the methods in Yuan et al. in [10], we constructed a matrix $U$ such that $\Omega = UU^T$, and where $U$ is a $9 \times 20$ matrix.
Then, the Ito stochastic differential model has the form,

$$d(X(t)) = f(X_1, X_2, X_3, X_4, X_5, X_6, X_7, X_8, X_9)dt + U \cdot dW(t)$$

with initial condition

$$X(0) = (X_1(0), X_2(0), X_3(0), X_4(0), X_5(0), X_6(0), X_7(0), X_8(0), X_9(0))^T$$

and a Wiener process, are along with the standard deviation $\sigma$

$$\begin{aligned} W(t) \quad = \quad & (W_1(t), W_2(t), W_3(t), W_4(t), W_5(t), W_6(t), W_7(t), W_8(t), W_9(t), W_{10}(t), \\ & W_{11}(t), W_{12}(t), W_{13}(t), W_{14}(t), W_{15}(t), W_{16}(t), W_{17}(t), W_{18}(t), W_{19}(t), W_{20}(t))^T \end{aligned}$$

The final SDE system of Model A is given by

$$\begin{aligned} dS &= (\pi_S - \mu_S S - aSV)dt \\ &\quad -\sqrt{\pi_S} dW_1 - \sqrt{\mu_S S} dW_2 - \sqrt{aSV} dW_3 \quad , \\ dI &= (aSV - (\mu_i + \mu_S)\, I)dt \\ &\quad +\sqrt{aSV} dW_3 - \sqrt{\mu_i I} dW_4 - \sqrt{\mu_S I} dW_5 \quad , \\ dV &= (\kappa\mu_i I - bSV - b_m S_m V - d_M MV - d_G GV)dt \\ &\quad +\sqrt{\kappa\mu_i I} dW_6 - \sqrt{V(bS + b_m S_m + d_M M + d_G G)} dW_7 \quad , \\ dS_m &= (\pi_m - \mu_S S_m - a_m S_m V)dt \\ &\quad -\sqrt{\pi_m} dW_8 - \sqrt{\mu_S S_m} dW_9 - \sqrt{a_m S_m V} dW_{10} \quad , \\ dP &= (a_m S_m V - \mu_s P - \mu_p P)dt \\ &\quad +\sqrt{a_m S_m V} dW_{10} - \sqrt{\mu_S P} dW_{11} - \sqrt{\mu_p P} dW_{12} \quad , \\ dM &= (\alpha_M P - \mu_M M - \gamma_M MV)dt \\ &\quad +\sqrt{\alpha_M P} dW_{13} - \sqrt{\mu_M M} dW_{14} - \sqrt{\gamma_M MV} dW_{15} \quad , \\ dG &= (\alpha_G P - \mu_G G - \gamma_G GV)dt \\ &\quad +\sqrt{\alpha_G P} dW_{16} - \sqrt{\mu_G G} dW_{17} - \sqrt{\gamma_G GV} dW_{18} \quad , \\ dC_M &= (\gamma_M MV - \mu_{C_M} C_M)dt \\ &\quad +\sqrt{\gamma_M MV} dW_{15} - \sqrt{\mu_{C_M} C_M} dW_{19} \quad , \\ dC_G &= (\gamma_G GV - \mu_{C_G} C_G)dt \\ &\quad +\sqrt{\gamma_G GV} dW_{18} - \sqrt{\mu_{C_G} C_G} dW_{20} \quad . \end{aligned} \tag{3}$$

Note that the construction of the SDE system for Model B and Model C follows the same steps as described in detail for Model A.

##### 2.1.2 Model B

The deterministic Model B is given by

$$\begin{aligned}
\frac{dS}{dt} &= \pi_S - \mu_S S - aSV \\
\frac{dI}{dt} &= aSV - (\mu_i + \mu_S)\, I \\
\frac{dV}{dt} &= \kappa\mu_i I - bSV - b_m S_m V - d_M MV - d_G GV \\
\frac{dS_m}{dt} &= \pi_m - \mu_S S_m - a_m S_m V \\
\frac{dP}{dt} &= a_m S_m V - (\mu_S + \mu_P)P \\
\frac{dM}{dt} &= \alpha_M P - \mu_M M - \gamma_M MV \\
\frac{dG}{dt} &= \alpha_G P - \gamma_G GV - \mu_G G + \alpha G_{sec} V \\
\frac{dC_M}{dt} &= \gamma_M MV - \mu_{C_M} C_M \\
\frac{dC_G}{dt} &= \gamma_G GV - \mu_{C_G} C_G \quad ,
\end{aligned} \tag{4}$$

and the state changes and probabilities for Model B are presented in Table S4.

Table S4: Possible changes of states and their probabilities for models B.

| Possible state change | Probability of state change |
|---|---|
| $(\Delta X)_1 = (1,0,0,0,0,0,0,0,0)^T$ | $P_1 = \pi_S \Delta t + O(\Delta t)$ |
| $(\Delta X)_2 = (-1,0,0,0,0,0,0,0,0)^T$ | $P_2 = \mu_S X_1 \Delta t + O(\Delta t)$ |
| $(\Delta X)_3 = (-1,1,0,0,0,0,0,0,0)^T$ | $P_3 = aX_1X_3\Delta t + O(\Delta t)$ |
| $(\Delta X)_4 = (0,-1,0,0,0,0,0,0,0)^T$ | $P_4 = \mu_i X_2 \Delta t + O(\Delta t)$ |
| $(\Delta X)_5 = (0,-1,0,0,0,0,0,0,0)^T$ | $P_5 = \mu_S X_2 \Delta t + O(\Delta t)$ |
| $(\Delta X)_6 = (1,0,0,0,0,0,0,0,0)^T$ | $P_6 = \kappa\mu_i X_2 \Delta t + O(\Delta t)$ |
| $(\Delta X)_7 = (0,0,-1,0,0,0,0,0,0)^T$ | $P_7 = X_3(bX_1 + b_m X_4 + d_M X_6 + d_G X_7)\Delta t + O(\Delta t)$ |
| $(\Delta X)_8 = (0,0,0,1,0,0,0,0,0)^T$ | $P_8 = \pi_m \Delta t + O(\Delta t)$ |
| $(\Delta X)_9 = (0,0,0,-1,0,0,0,0,0)^T$ | $P_9 = \mu_S X_4 \Delta t + O(\Delta t)$ |
| $(\Delta X)_{10} = (0,0,0,-1,1,0,0,0,0)^T$ | $P_{10} = a_m X_3 X_4 \Delta t + O(\Delta t)$ |
| $(\Delta X)_{11} = (0,0,0,0,-1,0,0,0,0)^T$ | $P_{11} = \mu_S X_5 \Delta t + O(\Delta t)$ |
| $(\Delta X)_{12} = (0,-1,0,0,0,0,0,0,0,0)^T$ | $P_{12} = \mu_p X_5 \Delta t + O(\Delta t)$ |
| $(\Delta X)_{13} = (0,0,0,0,0,1,0,0,0)^T$ | $P_{13} = \alpha_M X_5 \Delta t + O(\Delta t)$ |
| $(\Delta X)_{14} = (0,0,0,0,0,-1,0,0,0)^T$ | $P_{14} = \mu_M X_6 \Delta t + O(\Delta t)$ |
| $(\Delta X)_{15} = (0,0,0,0,0,-1,0,1,0)^T$ | $P_{15} = \gamma_M X_3 X_6 \Delta t + O(\Delta t)$ |
| $(\Delta X)_{16} = (0,0,0,0,0,0,1,0,0)^T$ | $P_{16} = \alpha_G X_5 \Delta t + O(\Delta t)$ |
| $(\Delta X)_{17} = (0,0,0,0,0,0,-1,0,0)^T$ | $P_{17} = \mu_G X_7 \Delta t + O(\Delta t)$ |
| $(\Delta X)_{18} = (0,0,0,0,0,0,-1,0,1)^T$ | $P_{18} = \gamma_G X_3 X_7 \Delta t + O(\Delta t)$ |
| $(\Delta X)_{19} = (0,0,0,0,0,0,1,0,0)^T$ | $P_{19} = \alpha_{G_{sec}} X_3 \Delta t + O(\Delta t)$ |
| $(\Delta X)_{20} = (0,0,0,0,0,0,0,-1,0)^T$ | $P_{20} = \mu_{CM} X_8 \Delta t + O(\Delta t)$ |
| $(\Delta X)_{21} = (0,0,0,0,0,0,0,0,-1)^T$ | $P_{21} = \mu_{CG} X_9 \Delta t + O(\Delta t)$ |
| $(\Delta X)_{22} = (0,0,0,0,0,0,0,0,0)^T$ | $P_{22} = 1 - \sum_{i=1}^{21} \Delta t + O(\Delta t)$ |

The final SDE system of Model B is given by

$$
\begin{aligned}
dS &= (\pi_S - \mu_S S - aSV)dt \\
&\quad -\sqrt{\pi_S}dW_1 - \sqrt{\mu_S S}dW_2 - \sqrt{aSV}dW_3 \quad , \\
dI &= (aSV - (\mu_i + \mu_S)\, I)dt \\
&\quad +\sqrt{aSV}dW_3 - \sqrt{\mu_i I}dW_4 - \sqrt{\mu_S I}dW_5 \quad , \\
dV &= (\kappa\mu_i I - bSV - b_m S_m V - d_M MV - d_G GV)dt \\
&\quad +\sqrt{\kappa\mu_i I}dW_6 - \sqrt{V(bS + b_m S_m + d_M M + d_G G)}dW_7 \quad , \\
dS_m &= (\pi_m - \mu_S S_m - a_m S_m V)dt \\
&\quad -\sqrt{\pi_m}dW_8 - \sqrt{\mu_S S_m}dW_9 - \sqrt{a_m S_m V}dW_{10} \quad , \\
dP &= (a_m S_m V - \mu_s P - \mu_p P)dt \\
&\quad +\sqrt{a_m S_m V}dW_{10} - \sqrt{\mu_S P}dW_{11} - \sqrt{\mu_p P}dW_{12} \quad , \\
dM &= (\alpha_M P - \mu_M M - \gamma_M MV)dt \\
&\quad +\sqrt{\alpha_M P}dW_{13} - \sqrt{\mu_M M}dW_{14} - \sqrt{\gamma_M MV}dW_{15} \quad , \\
dG &= (\alpha_G P - \mu_G G - \gamma_G GV + \alpha G_{sec} V)dt \\
&\quad +\sqrt{\alpha_G P}dW_{16} - \sqrt{\mu_G G}dW_{17} - \sqrt{\gamma_G GV}dW_{18} + \sqrt{\alpha G_{sec} V}dW_{19} \quad , \\
dC_M &= (\gamma_M MV - \mu_{C_M} C_M)dt \\
&\quad +\sqrt{\gamma_M MV}dW_{15} - \sqrt{\mu_{C_M} C_M}dW_{20} \quad , \\
dC_G &= (\gamma_G GV - \mu_{C_G} C_G)dt \\
&\quad +\sqrt{\gamma_G GV}dW_{18} - \sqrt{\mu_{C_G} C_G}dW_{21} \quad .
\end{aligned} \tag{5}
$$

##### 2.1.3 Model C

The deterministic Model C is given by

$$\begin{aligned}
\frac{dS}{dt} &= \pi_S - \mu_S S - aSV - a_{ADE}SC_G \\
\frac{dI}{dt} &= aSV - (\mu_i + \mu_S)\,I + a_{ADE}SC_G \\
\frac{dV}{dt} &= \kappa\mu_i I - bSV - b_m S_m V - d_M MV - d_G GV \\
\frac{dS_m}{dt} &= \pi_m - \mu_S S_m - a_m S_m V \\
\frac{dP}{dt} &= a_m S_m V - (\mu_S + \mu_P)P \\
\frac{dM}{dt} &= \alpha_M P - \mu_M M - \gamma_M MV \\
\frac{dG}{dt} &= \alpha_G P - \gamma_G GV - \mu_G G + \alpha G_{sec} V \\
\frac{dC_M}{dt} &= \gamma_M MV - \mu_{C_M} C_M \\
\frac{dC_G}{dt} &= \gamma_G GV - \mu_{C_G} C_G - b_{ADE}SC_G
\end{aligned} \tag{6}$$

and the state changes and their probabilities are presented in Table S5.

Table S5: Possible changes of states and their probabilities for models C.

| Possible state change | Probability of state change |
|---|---|
| $(\Delta X)_1 = (1,0,0,0,0,0,0,0,0)^T$ | $P_1 = \pi_S \Delta t + O(\Delta t)$ |
| $(\Delta X)_2 = (-1,0,0,0,0,0,0,0,0)^T$ | $P_2 = \mu_S X_1 \Delta t + O(\Delta t)$ |
| $(\Delta X)_3 = (-1,1,0,0,0,0,0,0,0)^T$ | $P_3 = aX_1X_3\Delta t + O(\Delta t)$ |
| $(\Delta X)_4 = (-1,1,0,0,0,0,0,0,0)^T$ | $P_4 = a_{ADE}X_1X_9\Delta t + O(\Delta t)$ |
| $(\Delta X)_5 = (0,-1,0,0,0,0,0,0,0)^T$ | $P_5 = \mu_i X_2 \Delta t + O(\Delta t)$ |
| $(\Delta X)_6 = (0,-1,0,0,0,0,0,0,0)^T$ | $P_6 = \mu_S X_2 \Delta t + O(\Delta t)$ |
| $(\Delta X)_7 = (1,0,0,0,0,0,0,0,0)^T$ | $P_7 = \kappa\mu_i X_2 \Delta t + O(\Delta t)$ |
| $(\Delta X)_8 = (0,0,-1,0,0,0,0,0,0)^T$ | $P_8 = X_3(bX_1 + b_mX_4 + d_MX_6 + d_GX_7)\Delta t + O(\Delta t)$ |
| $(\Delta X)_9 = (0,0,0,1,0,0,0,0,0)^T$ | $P_9 = \pi_m \Delta t + O(\Delta t)$ |
| $(\Delta X)_{10} = (0,0,0,-1,0,0,0,0,0)^T$ | $P_{10} = \mu_S X_4 \Delta t + O(\Delta t)$ |
| $(\Delta X)_{11} = (0,0,0,-1,1,0,0,0,0)^T$ | $P_{11} = a_m X_3 X_4 \Delta t + O(\Delta t)$ |
| $(\Delta X)_{12} = (0,0,0,0,-1,0,0,0,0)^T$ | $P_{12} = \mu_S X_5 \Delta t + O(\Delta t)$ |
| $(\Delta X)_{13} = (0,-1,0,0,0,0,0,0,0)^T$ | $P_{13} = \mu_p X_5 \Delta t + O(\Delta t)$ |
| $(\Delta X)_{14} = (0,0,0,0,0,1,0,0,0)^T$ | $P_{14} = \alpha_M X_5 \Delta t + O(\Delta t)$ |
| $(\Delta X)_{15} = (0,0,0,0,0,-1,0,0,0)^T$ | $P_{15} = \mu_M X_6 \Delta t + O(\Delta t)$ |
| $(\Delta X)_{16} = (0,0,0,0,0,-1,0,1,0)^T$ | $P_{16} = \gamma_M X_3 X_6 \Delta t + O(\Delta t)$ |
| $(\Delta X)_{17} = (0,0,0,0,0,0,1,0,0)^T$ | $P_{17} = \alpha_G X_5 \Delta t + O(\Delta t)$ |
| $(\Delta X)_{18} = (0,0,0,0,0,0,-1,0,0)^T$ | $P_{18} = \mu_G X_7 \Delta t + O(\Delta t)$ |
| $(\Delta X)_{19} = (0,0,0,0,0,0,-1,0,1)^T$ | $P_{19} = \gamma_G X_3 X_7 \Delta t + O(\Delta t)$ |
| $(\Delta X)_{20} = (0,0,0,0,0,0,1,0,0)^T$ | $P_{20} = \alpha_{G_{sec}} X_3 \Delta t + O(\Delta t)$ |
| $(\Delta X)_{21} = (0,0,0,0,0,0,0,-1,0)^T$ | $P_{21} = \mu_{CM} X_8 \Delta t + O(\Delta t)$ |
| $(\Delta X)_{22} = (0,0,0,0,0,0,0,0,-1)^T$ | $P_{22} = \mu_{CG} X_9 \Delta t + O(\Delta t)$ |
| $(\Delta X)_{23} = (0,0,0,0,0,0,0,0,-1)^T$ | $P_{23} = b_{ADE} X_9 \Delta t + O(\Delta t)$ |
| $(\Delta X)_{24} = (0,0,0,0,0,0,0,0,0)^T$ | $P_{24} = 1 - \sum_{i=1}^{23} \Delta t + O(\Delta t)$ |

The final SDE system of Model C is given by

$$
\begin{aligned}
dS &= (\pi_S - \mu_S S - aSV - a_{ADE}SC_G)dt \\
&\quad -\sqrt{\pi_S}dW_1 - \sqrt{\mu_S S}dW_2 - \sqrt{aSV}dW_3 - \sqrt{a_{ADE}SC_G}dW_4 \quad , \\
dI &= (aSV - (\mu_i + \mu_S)\, I + a_{ADE}SC_G)dt \\
&\quad +\sqrt{aSV}dW_3 + \sqrt{a_{ADE}SC_G}dW_4 - \sqrt{\mu_i I}dW_5 - \sqrt{\mu_S I}dW_6 \quad , \\
dV &= (\kappa\mu_i I - bSV - b_m S_m V - d_M MV - d_G GV)dt \\
&\quad +\sqrt{\kappa\mu_i I}dW_7 - \sqrt{V(bS + b_m S_m + d_M M + d_G G)}dW_8 \quad , \\
dS_m &= (\pi_m - \mu_S S_m - a_m S_m V)dt \\
&\quad -\sqrt{\pi_m}dW_9 - \sqrt{\mu_S S_m}dW_{10} - \sqrt{a_m S_m V}dW_{11} \quad , \\
dP &= (a_m S_m V - \mu_s P - \mu_p P)dt \\
&\quad +\sqrt{a_m S_m V}dW_{11} - \sqrt{\mu_S P}dW_{12} - \sqrt{\mu_p P}dW_{13} \quad , \\
dM &= (\alpha_M P - \mu_M M - \gamma_M MV)dt \\
&\quad +\sqrt{\alpha_M P}dW_{14} - \sqrt{\mu_M M}dW_{15} - \sqrt{\gamma_M MV}dW_{16} \quad , \\
dG &= (\alpha_G P - \mu_G G - \gamma_G GV + \alpha G_{sec}V)dt \\
&\quad +\sqrt{\alpha_G P}dW_{17} - \sqrt{\mu_G G}dW_{18} - \sqrt{\gamma_G GV}dW_{19} + \sqrt{\alpha G_{sec}V}dW_{20} \quad , \\
dC_M &= (\gamma_M MV - \mu_{C_M} C_M)dt \\
&\quad +\sqrt{\gamma_M MV}dW_{16} - \sqrt{\mu_{C_M} C_M}dW_{21} \quad , \\
dC_G &= (\gamma_G GV - \mu_{C_G} C_G - b_{ADE}SC_G)dt \\
&\quad +\sqrt{\gamma_G GV}dW_{19} - \sqrt{\mu_{C_G} C_G}dW_{22} - \sqrt{b_{ADE}SC_G}dW_{23} \quad .
\end{aligned}
\tag{7}
$$

#### 2.2 Modeling simulations and robustness analysis

Model simulations were conducted using MATLAB software. For the deterministic system, the simulations were obtained using the ode45 and ode15s methods, and for the stochastic simulations, the Euler-Maruyama method [9] was used. The baseline parameter values used for modeling simulations are obtained from the literature and listed in Table S1.

The SDE dynamical behaviour for each model variable is shown in Figure S6. The results obtained for Model A, Model B and Model C are shown in Figures S6-S8.

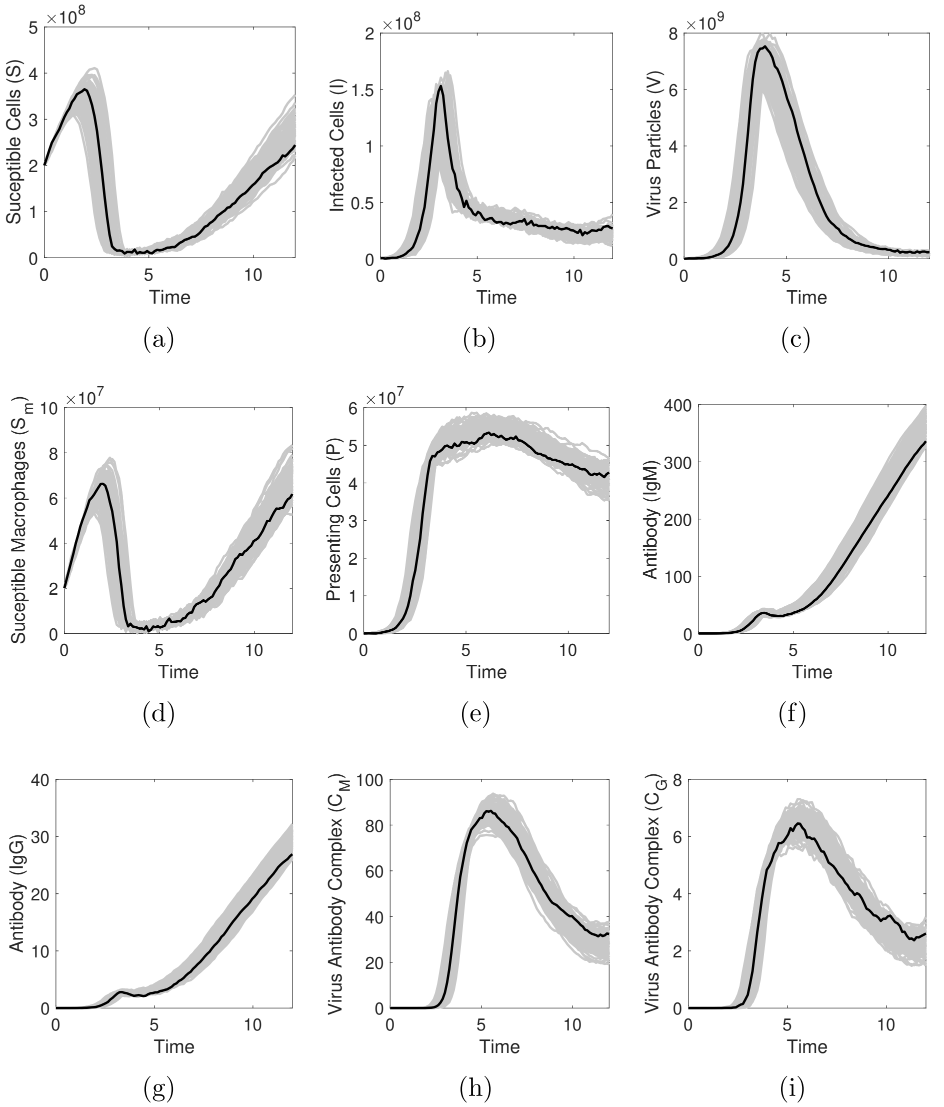


Figure S6: Model A. The mean (black lines) of 100 stochastic realizations (gray lines).

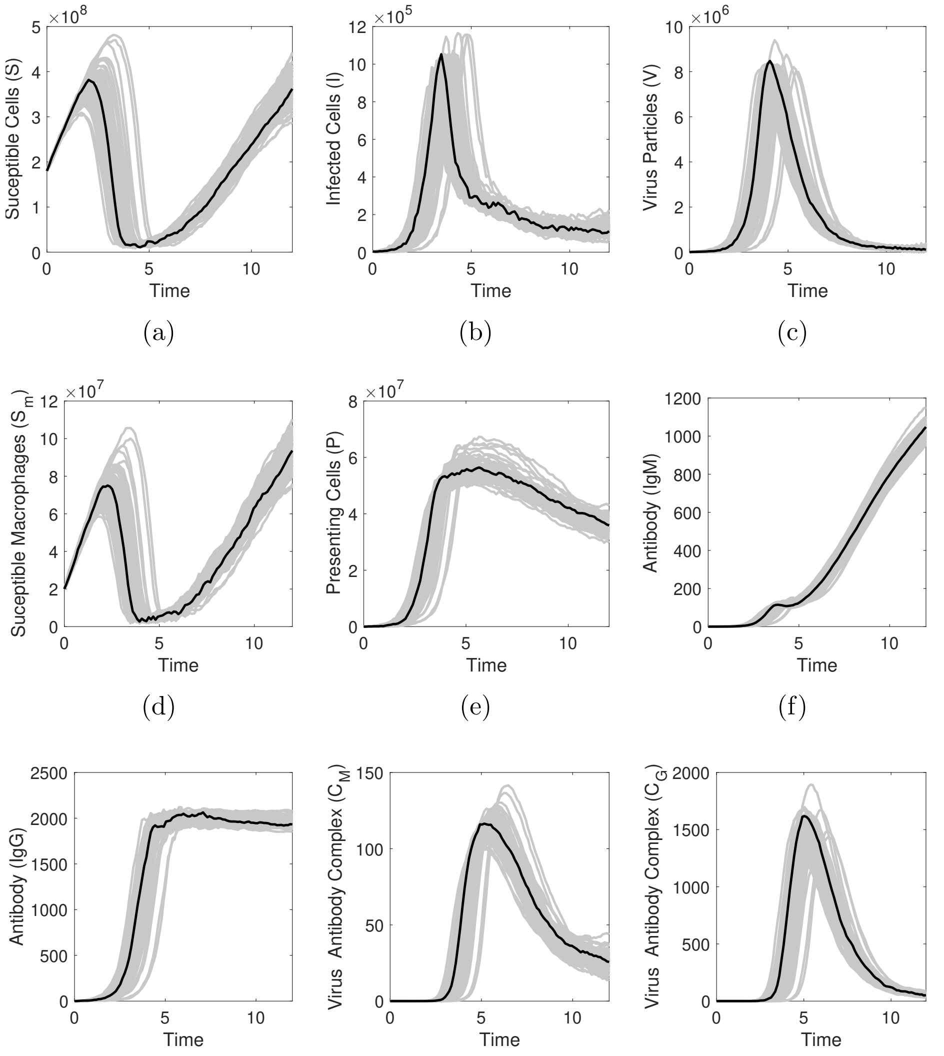


(a) (b) (c)

(d) (e) (f)

(g) (h) (i)

Figure S7: Model B. The mean (black lines) of 100 stochastic realizations (gray lines)

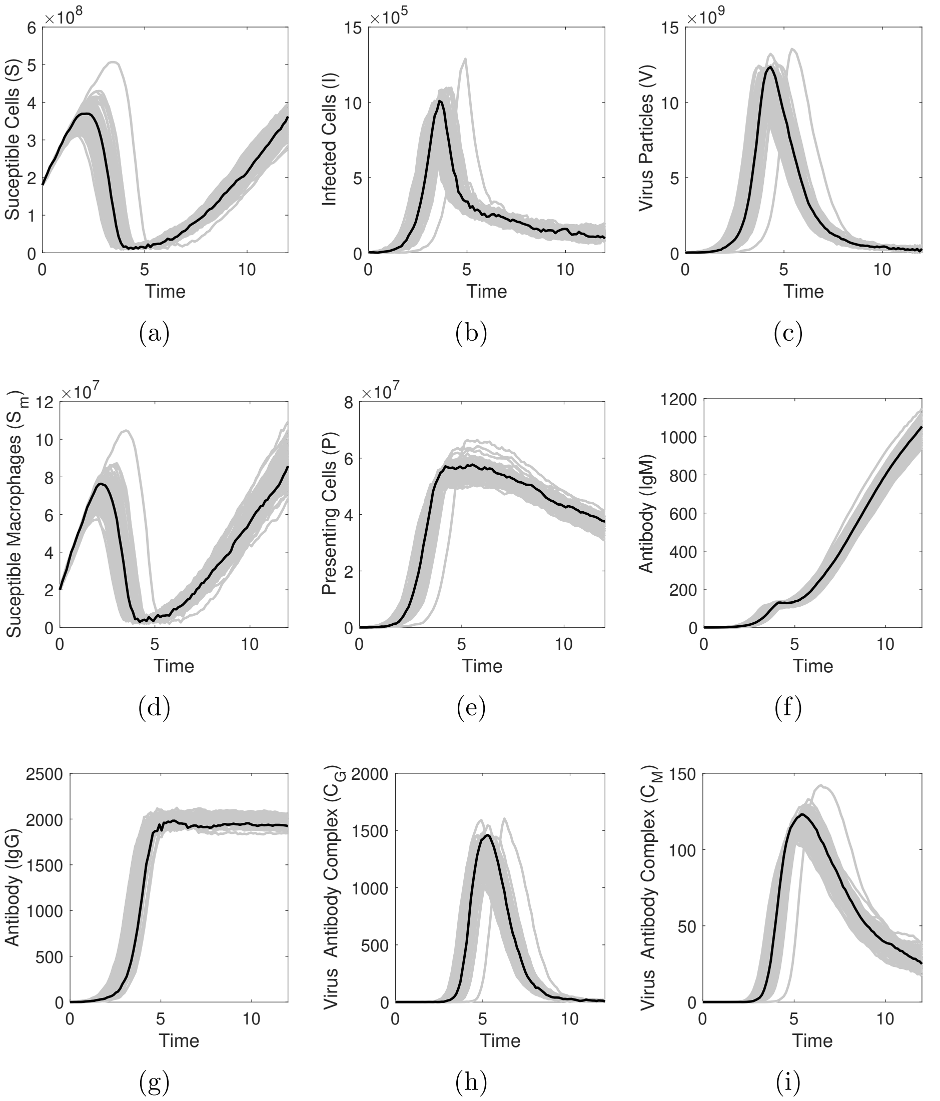


Figure S8: Model C. The mean (black lines) of 100 stochastic realizations (gray lines)

To assess the robustness of our findings, we have compared the mean of 100 stochastic realizations with the mean field solution (deterministic system) for each one of the model variables. The results obtained for Model A, Model B and Model C are shown in Figures S9-S11.

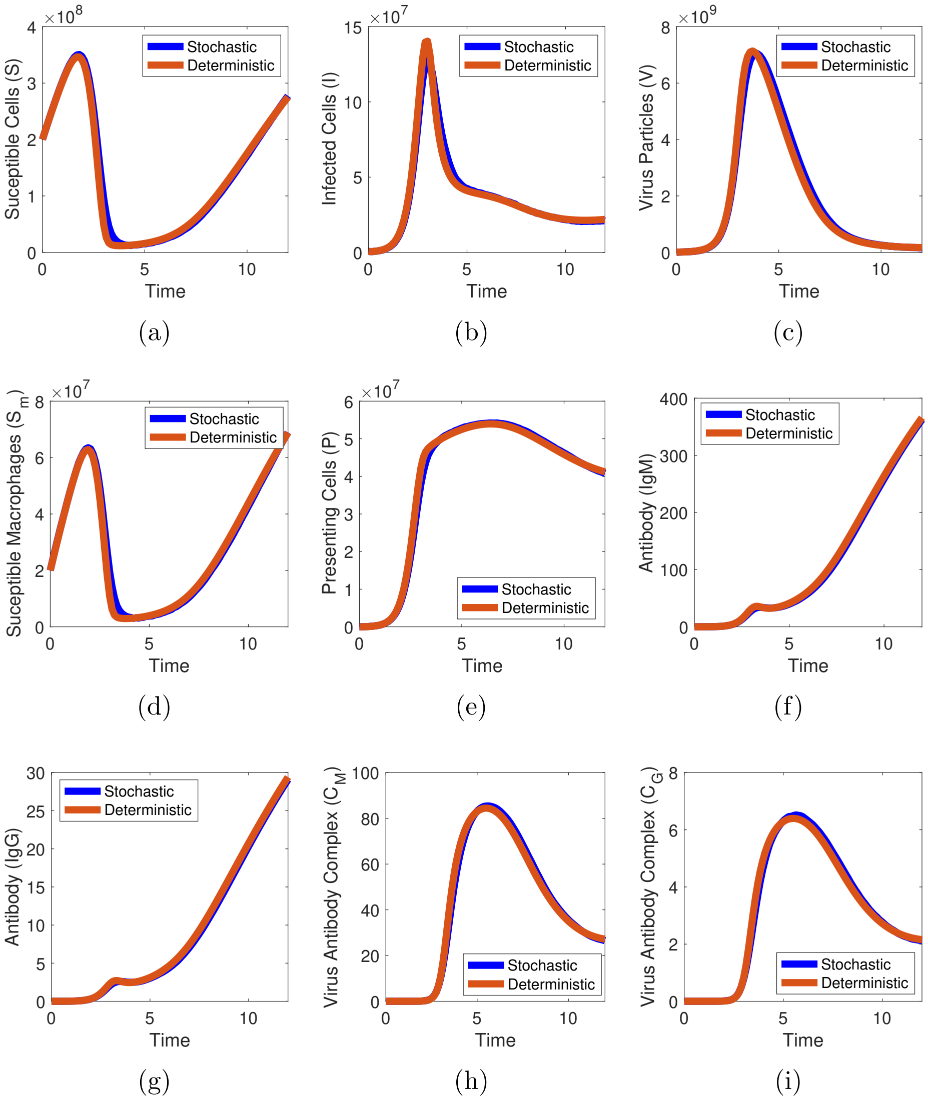


Figure S9: Model A. Comparison of the mean of stochastic realizations (blue lines) with the deterministic solution (red lines).

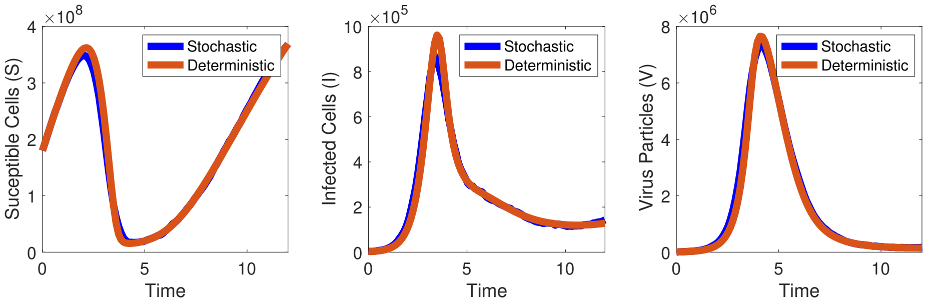


(a) (b) (c)

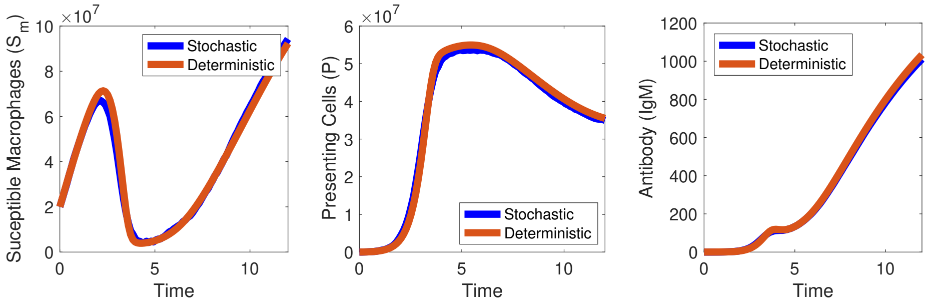


(d) (e) (f)

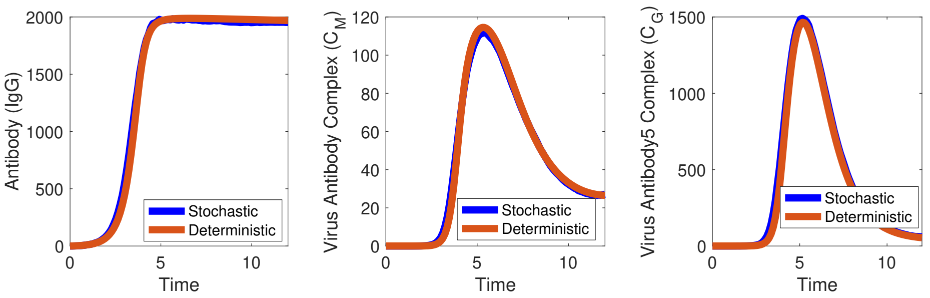


(g) (h) (i)

Figure S10: Model B. Comparison of the mean of stochastic realizations (blue lines) with the deterministic solution (red lines).

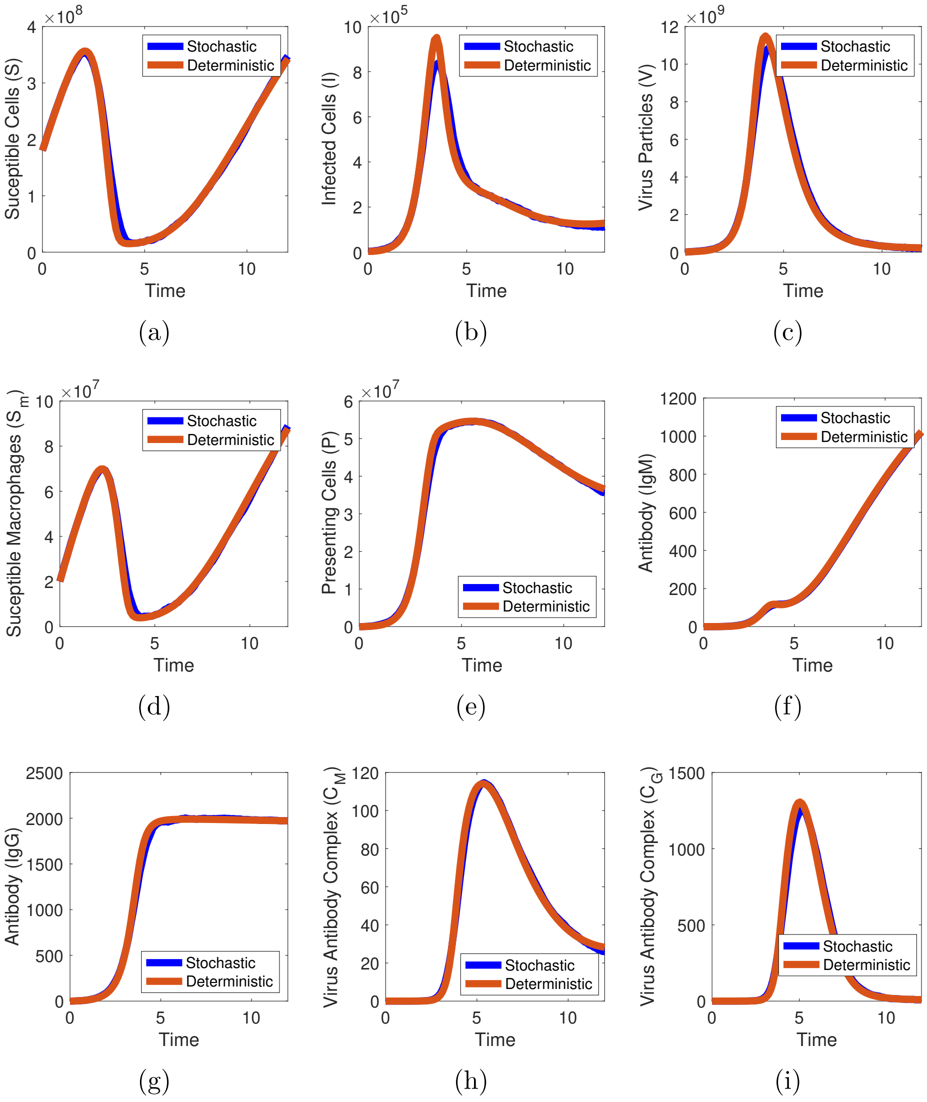


Figure S11: Model C. Comparison of the mean of stochastic realizations (blue lines) with the deterministic solution (red lines).

### References

[1] Nguyen NM, Tran CNB, Phung LK, Duong KTH, Huynh HlA, Farrar J, et al. A randomized, double-blind placebo controlled trial of balapiravir, a polymerase inhibitor, in adult dengue patients. The Journal of infectious diseases. 2013;207(9):1442-50.

[2] Clapham HE, Quyen TH, Kien DTH, Dorigatti I, Simmons CP, Ferguson NM. Modelling virus and antibody dynamics during dengue virus infection suggests a role for antibody in virus clearance. PLoS computational biology. 2016;12(5):e1004951.

[3] Clapham HE, Tricou V, Van Vinh Chau N, Simmons CP, Ferguson NM. Within-host viral dynamics of dengue serotype 1 infection. Journal of the Royal Society Interface. 2014;11(96):20140094.

[4] Aguiar M, Anam V, Blyuss KB, Estadilla CDS, Guerrero BV, Knopoff D, et al. Mathematical models for dengue fever epidemiology: a 10-year systematic review. Physics of Life Reviews. 2022.

[5] Ben-Shachar R, Koelle K. Minimal within-host dengue models highlight the specific roles of the immune response in primary and secondary dengue infections. Journal of the Royal Society Interface. 2015;12(103):20140886.

[6] Cologna R, Rico-Hesse R. American genotype structures decrease dengue virus output from human monocytes and dendritic cells. Journal of virology. 2003;77(7):3929-38.

[7] Wahala WM, De Silva AM. The human antibody response to dengue virus infection. Viruses. 2011;3(12):2374-95.

[8] Edward, J Allen L, J S Allen A, Arciniega P, Greenwood E. Construction of Equivalent Stochastic Differential Equation Models. Stochastic Analysis and Applications. 2008 march;26(2):274-97. Available from: `http://https://www.tandfonline.com/doi/abs/10.1080/07362990701857129`.

[9] E P, Kloeden E, Platen. Numerical Solution of Stochastic Differential Equations. In: Stochastic Modelling and Applied Probability. Springer Berlin Heidelberg; 1992. p. 95-463.

[10] Yuan Y, Allen LJ. Stochastic models for virus and immune system dynamics. Mathematical biosciences. 2011;234(2):84-94.
